# Genetic determinants of daytime napping and effects on cardiometabolic health

**DOI:** 10.1101/2020.06.12.20129858

**Authors:** Hassan S. Dashti, Iyas Daghlas, Jacqueline M. Lane, Yunru Huang, Miriam S. Udler, Heming Wang, Hanna M. Ollila, Samuel E. Jones, Jaegil Kim, Andrew R. Wood, 23andMe Research Team, Michael N. Weedon, Stella Aslibekyan, Marta Garaulet, Richa Saxena

## Abstract

Daytime napping is a common, heritable behavior, but its genetic basis and causal relationship with cardiometabolic health remains unclear. Here, we performed a genome-wide association study of self-reported daytime napping in the UK Biobank (*n*=452,633) and identified 123 loci of which 60 replicated in 23andMe research participants (*n*=541,333). Findings included missense variants in established drug targets (*HCRTR1, HCRTR2*), genes with roles in arousal (*TRPC6, PNOC*), and genes suggesting an obesity-hypersomnolence pathway (*PNOC, PATJ*). Signals were concordant with accelerometer-measured daytime inactivity duration and 33 signals colocalized with signals for other sleep phenotypes. Cluster analysis identified 3 clusters suggesting distinct nap-promoting mechanisms with heterogeneous associations with cardiometabolic outcomes. Mendelian randomization showed potential causal links between more frequent daytime napping and higher systolic blood pressure, diastolic blood pressure, and waist circumference.

---

Naps are short daytime sleep episodes that are evolutionarily conserved across diverse diurnal species ranging from flies^1^ to polyphasic mammals^2^. In human adults, daytime napping is highly prevalent in Mediterranean cultures and is also common in non-Mediterranean countries including the United States^3^. In modern society, napping is encouraged in sleep-deprived populations, such as night shift workers^4^ and airline pilots^5^, to acutely improve performance and alertness. Although an acute benefit of napping on increased arousal in the setting of sleep deprivation is well-established^6^, the long-term effects of habitual napping on chronic disease remains controversial. Indeed, cross-sectional studies have provided conflicting evidence effects of habitual napping on cognition, blood pressure, obesity, metabolic traits, and mortality^7–13^. As napping behavior may be confounded by inadequate nighttime sleep or underlying poor health^14,15^, causal inference in these observational studies is limited.

Genetic variation constitutes an important contributor to inter-individual differences in napping preference. A twin study estimated heritability of self-reported napping and objective daytime sleep duration at 65% and 61%, respectively, demonstrating heritability similar or even higher than those found for other sleep traits such as nighttime sleep duration or timing^16^. Indeed, up to 7 genetic loci for daytime napping have been discovered in genome-wide association study (GWAS) of self-reported napping or related accelerometer-derived sleep measures^17–19^. Discovery of additional genetic loci could reveal novel biological pathways regulating sleep, elucidate genetic links with other sleep and metabolic traits, and clarify the potential causal effects of habitual napping on cardiometabolic disease. We therefore leveraged the full UK Biobank dataset of European ancestry, including related individuals (*n* =452,633), and an independent replication sample from 23andMe research participants of European ancestry (*n* =541,333), to define the genetic architecture of daytime napping and to assess links with other sleep and cardiometabolic traits.

## Results

Among UK Biobank participants of European ancestry (*n* =452,633), 38.2% and 5.3% of participants reported *sometimes* and *always* napping, respectively (**Supplementary Table 1**). Overall, participants reporting always napping were more likely to be older males, report longer 24h sleep duration, have higher body mass index (BMI), waist circumference, systolic and diastolic blood pressures, have a higher Townsend deprivation index (i.e., greater degree of socio-economic deprivation), report being current smokers, unemployed or retired, and shift workers (all *P* <0.001; **Supplementary Table 1**).

### Discovery, validation, and replication of 123 genetic loci for daytime napping in UK Biobank and 23andMe

We conducted GWAS using 13,304,133 high-quality imputed genetic variants across 452,633 participants. We identified 123 distinct loci, with (*P* <5×10^−8^; **Figure 1a, Supplementary Table 2, Supplementary Figure 1a**) genome-wide SNP-based heritability estimated at 11.9% (standard error =0.1%). The 123 loci explained 1.1% of the variance in daytime napping. The LD score regression intercept was 1.04 and therefore did not indicate uncontrolled confounding. Effects were largely consistent in GWAS restricted to 338,764 participants self-reporting *excellent* or *good* overall health (**Supplementary Table 1,2**). As higher BMI is associated with more frequent napping^20^, we conducted a GWAS adjusting for BMI and found that 110 of the 123 loci retained genome-wide significance (**Supplementary Table 2**). In addition, we found no evidence of sexual dimorphism in the genetic determinants of daytime napping behavior^21^ as indicated by the lack of statistical heterogeneity by sex at any of the lead loci (all *P* >0.005) (**Supplementary Table 2**) and a genome-wide genetic correlation (*r*_g_) of male and female stratified GWAS of 0.94 (standard error =0.03). In addition, 5 of 7 loci for daytime napping reported in earlier GWAS in a subset of unrelated UK Biobank participants of European ancestry (*n* =386,577)^18^ retained significance in our analyses (**Supplementary Table 3**). However, none of the suggestive loci reported in GWAS of accelerometer-derived phenotypes related to napping in the UK Biobank (*n* =85,670)^19^ and LIFE Adult Study (*n* =956)^17^ showed evidence of association in the current analysis.

**Figure 1.**
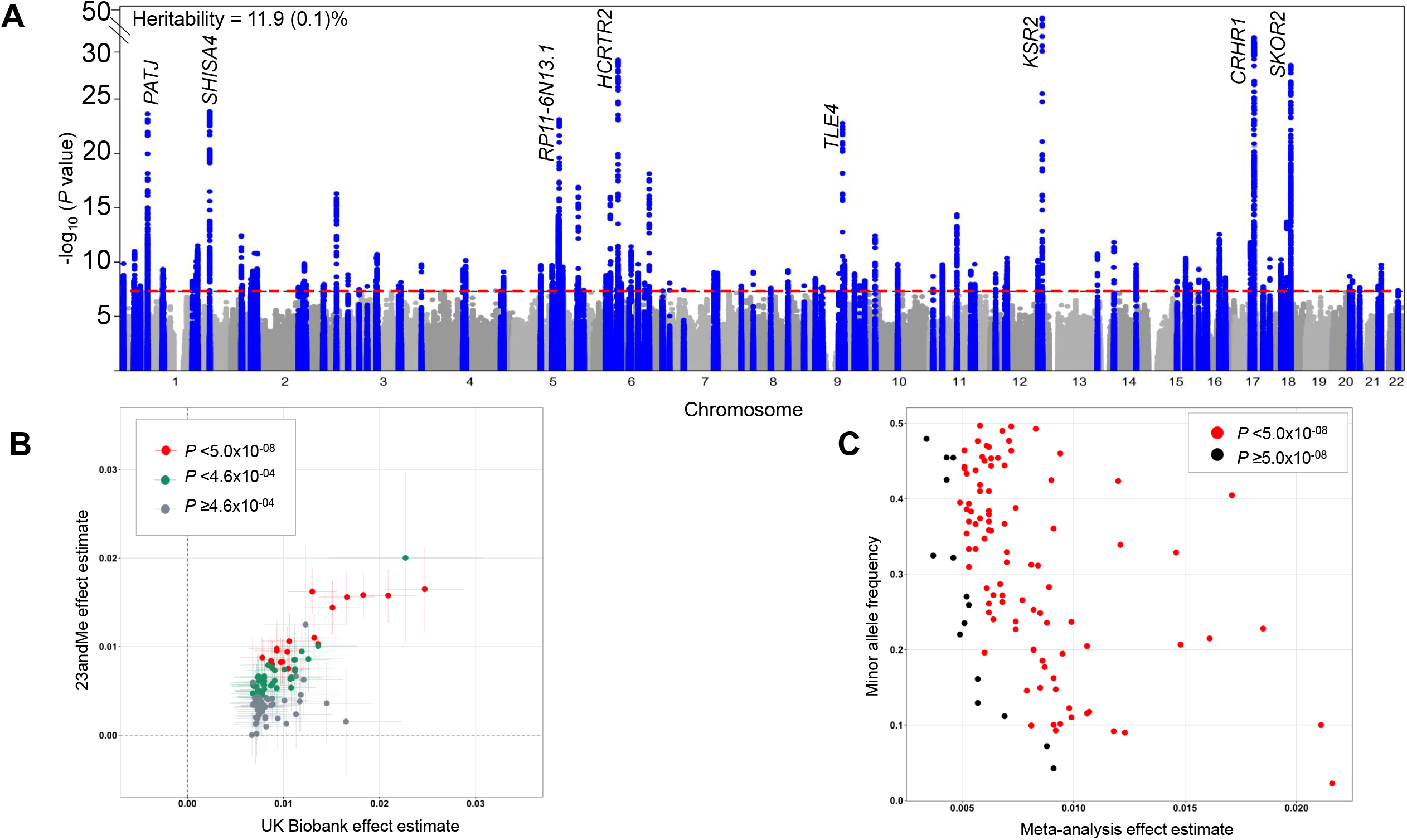
Plots for genome-wide association analysis results for daytime napping in the UK Biobank *(n* =452,633) and replication in 23andMe (*n* =541,333). A. Manhattan plot of daytime napping genome-wide association study in the UK Biobank (*n* =452,633). Plot shows the −log_10_*P* values (y-axis) for all genotyped and imputed single-nucleotide polymorphisms (SNPs) passing quality control plotted by chromosome (x-axis). Blue peaks represent genome-wide significant loci. Horizontal red line denotes genome-wide significance (*P* =5×10^−8^). Top 8 loci are annotated with nearest gene. B. Comparison of daytime napping signals’ effect estimates for UK Biobank (*n* =452,633) and 23andMe (*n* =541,333). Error bars represent the 95% confidence intervals for each effect estimate. C. Effect estimates of daytime napping signals from UK Biobank and 23andMe meta-analysis (*n* =993,966) plotted against minor allele frequency.

We tested for independent replication of lead loci using data from 23andMe, Inc., a personal genetics company, where 541,333 research participants of European ancestry (43.0% *sometimes* and 7.6% *always* napping) also provided data on the frequency of daytime napping (**Supplementary Table 4**). We replicated 60 of 109 tested loci (*P* <4.07×10^−4^), of which 18 loci were genome-wide significant. All 109 loci showed consistent direction of effect with the UK Biobank (*P*_binomial_ =3.21×10^−8^) (**Figure 1b, Supplementary Table 5**). In inverse-variance fixed-effects weighted meta-analysis of UK Biobank and 23andMe (total *n* =993,966), 94 of the 109 remained genome-wide significant (**Figure 1c, Supplementary Table 5**).

Using 7-day wrist accelerometry data obtained in 85,499 participants of European ancestry in the UK Biobank >2 years after baseline assessment, we compared effect estimates at the 123 loci for self-reported daytime napping with effect estimates for accelerometer-derived daytime inactivity duration^19^, an objective measure corresponding to self-reported daytime napping. Estimates of 90 variants were directionally concordant (*P*_binomial_ =2.74×10^−7^) and variants at *ASCL4* and *SNAP91* were strongly associated with higher duration of daytime inactivity (*P*_adj_ <0.05) (**Supplementary Table 6**). We further quantified the impact of daytime napping on daytime inactivity duration using a polygenic score comprised of lead variants at all 123 loci. A category increase in frequency of daytime napping was associated with 18.9 minutes (95% confidence interval =13.6, 24.2; *P* =4.21×10^−12^) longer duration of daytime inactivity and had no effect on other accelerometer-derived sleep duration, timing or quality phenotypes **(Supplementary Table 7)**.

### Napping genetic variants share causal variants with other sleep phenotypes and lie near known genes that regulate arousal

Individual daytime napping loci also associated with self-reported sleep traits^22–25^ and accelerometer-derived sleep measures^19^ (**Supplementary Table 6, 8**). However, lead variants at 26 of the 123 loci showed no statistical evidence for association with previously studied sleep traits in the UK Biobank (*P*_adj_ >0.05), suggesting these variants might reflect mechanisms specific to daytime napping (**Supplementary Table 8**). Overlap was also supported by cross-trait LD score regression^26^ where we observed strongest evidence for a shared genetic basis with daytime sleepiness (*r*_g_ =0.70, *P* =7.94×10^−373^) and long sleep duration (*r*_g_ =0.42, *P* =1.94×10^−64^), and moderate to weak correlations with other sleep duration, timing and quality phenotypes (**Supplementary Table 9**).

Several genetic variants for daytime napping were located in or near genes with known effects on sleep/wake regulation. Thus, to gain insights into putative causal variants driving daytime napping and sleep/wake biology, we integrated results from functional annotation, fine-mapping, multi-trait and eQTL colocalization analyses; for each analysis we report a posterior probability for a shared causal variant in the association signal) (**Supplementary Table 10-13**). Functional annotation of all variants identified an enrichment of variants in intronic (46.2%) and intergenic (31.5%) regions, suggesting that non-coding gene regulatory mechanisms may underlie napping as they do for many other complex traits (**Supplementary Figure 1b**).

In order to identify association signals with evidence for shared causal variants with other sleep traits, we performed multi-trait colocalization analyses^27^ of daytime napping loci across six self-reported sleep traits (daytime sleepiness, sleep duration, insomnia, snoring, chronotype, and ease of awakening) and identified 33 shared signals (of which 25 corresponded to a genome-wide significant daytime napping locus) for which causal variants were prioritized (**Supplementary Table 10**).

These analyses prioritized putatively causal SNPs genes at several loci which may form hypotheses for experimental follow-up. First, missense variants were identified in components of the wake-promoting orexin/hypocretin neuropeptide signaling pathway: i) in a transmembrane helical domain of *HCRTR2* [I308V; rs2653349; A effect allele frequency (EAF)=0.21; associated with more frequent daytime napping, morning preference and ease of awakening, posterior probability of colocalization (pp)=0.98] reflecting putative loss of function, ii) in a cytoplasmic domain of *HCRTR1* [I408V; rs2271933, r^2^ 0.98 with lead rs6663012 variant; A EAF=0.38] associated with more frequent daytime napping again reflecting putative loss of function, and iii) a cytoplasmic domain of *TRPC6* [P15S; rs3802829, *r*^2^ =0.98 with lead rs11224896 variant; G EAF =0.11; associated with more napping and longer sleep duration; pp=0.80], which encodes a subunit for transient receptor channels that maintains hypocretin/orexin neurons in a depolarized state^28^. Although an intronic lead variant in *HCRTR2* was previously reported in GWAS of daytime sleepiness^23^ (rs3122170, *r*^2^=0.29 with lead napping variant rs2653349), the colocalization cluster excluded the daytime sleepiness phenotype, suggesting that the napping signal is driven by a distinct causal variant (**Supplementary Table 11**). As further evidence that two distinct signals drive the daytime sleepiness and daytime napping associations, we conducted conditional analysis using GCTA COJO adjusting the daytime napping association for the lead daytime napping variant (rs2653349), and observed attenuation in the association for the lead daytime sleepiness variant in *HCRTR2* (rs3122170) (*P* value from 4.60×10^−18^ to 4.56×10^−3^).

Second, colocalization revealed a set of variants with effects on napping, daytime sleepiness, and BMI, suggesting a novel obesity-hypersomnolence pathway. An intronic candidate causal variant in *PNOC* [rs351776; C EAF =0.55], associated with more frequent napping, more daytime sleepiness, and higher BMI. *PNOC* encodes a preproprotein that is proteolytically processed to generate the nociceptin neuropeptide, which opposes the effects of hypocretin to reduce arousal and spontaneous activity in zebrafish^29,30^. The colocalization of daytime napping with BMI at this locus is consistent with known pleiotropic effects of *PNOC* in feeding behavior^31^ (pp=0.93; **Supplementary Table 12**). The known^32^ missense variant in *PATJ* [rs12140153; G1543V; G EAF 0.90] has a stronger association with daytime napping than any previously studied sleep phenotypes (**Supplementary Table 8**), and is likely to be a shared causal variant with daytime sleepiness, chronotype, and BMI (pp=0.81; **Supplementary Tables 11-12**).

Third, colocalization analyses refined genetic effects previously described at the *KSR2* locus implicated in ERK/EGFR signaling^23^, a pathway with an established causal role in sleep regulation in *C. elegans, Drosophila*, and zebrafish^33,34^. This included an intronic variant in *KSR2*, a gene that encodes a scaffold protein for ERK cascade signaling^35^ (rs1846644; T allele AF=0.40 signal associated with higher napping frequency, longer sleep duration and daytime sleepiness, with 0.55 PICS probability of being the causal variant).

Fourth, several genetic variants were prioritized at or near genes a) coding for proteins constituting or interacting with potassium channels [rs77154532 (*KCHN8*), rs10875606 (*KCTD16*)], b) involved in glutamate transmission [rs60920123 (*GRIN2A*), rs2284015 (*CACNG2*)], and c) involved in lipid metabolism [rs12451365 (*ACACA*), rs174541 (*FADS1/FADS2*)].

Fifth, we found evidence of association for variants in *PRRC2C*, one of three orthologs of the *Drosophila* nocte gene^36^. Nocte targets clock neurons to synchronize molecular and behavioral rhythms to temperature cycles and influences siesta sleep in flies. Of note, we observed no gene-by-season (a proxy for ambient temperature) statistical interaction at this and any other loci (**Supplementary Table 2**).

Sixth, integration of gene expression data from the frontal cortex in the GTEx data release v7^37^ (*n* =129), the brain tissue predominantly enriched for daytime napping signals, showed that daytime napping variants at *FADS1* associated with increased expression of *FADS1* (pp=0.89) and at *ECE2* associated with increased expression of *ECE2* (endothelin converting enzyme; pp=0.99) (**Figure 2a,b**; **Supplementary Table 13**). Another lead variant is near *FNDC5* (rs2786547), a gene coding for irisin, a muscle-derived hormone with putative effects on expression of sleep-regulating neuropeptides^38^. We found strong evidence for colocalization of the daytime napping signal with gene expression in skeletal muscle in the GTEx data release v7^37^ (*n* =706, pp=0.93), with higher gene expression relating to reduced daytime napping frequency (**Supplementary Figure 2)**. This suggests a potentially novel sleep-regulating mechanism outside of the central nervous system.

**Figure 2.**
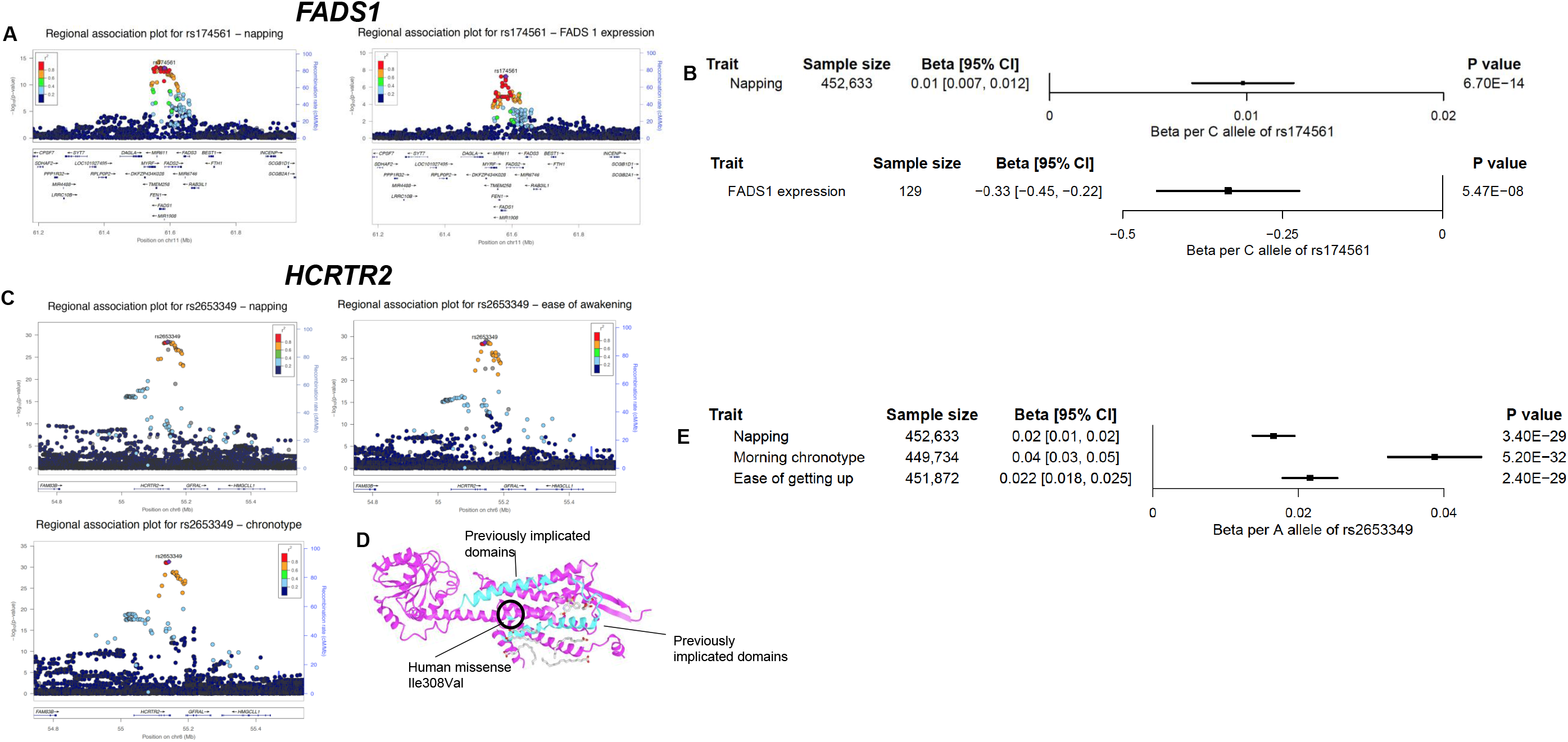
Colocalization analysis reveals a shared causal variant reducing FADS1 gene expression in the frontal cortex and increasing napping liability, and a shared causal missense variant in HCRTR2 influencing daytime napping, chronotype, and ease of awakening. A. Regional association plots for daytime napping and FADS1 gene expression at rs174561 and variants within 400kb on chromosome 11. The y-axis shows the -log_10_ P value for each variant in the region, and the x-axis shows the genomic position. Each variant is represented by a filled circle, with the rs174561 variant colored purple, and nearby variants colored according to degree of linkage disequilibrium (*r*^2^) with rs174561. The lower panel shows genes located in the displayed region and the blue line corresponds to the recombination rate. B. Forest plot of associations between the C allele of genetic variant rs174561 in FADS1 with daytime napping and gene expression of FADS1 in the frontal cortex. Units of daytime napping reflect an increase on the ordinal scale of the trait, and gene expression is in standard deviation units. Black box indicates the effect estimate and lines represent 95% confidence intervals. C. Regional association plot for colocalized sleep phenotypes at rs2653349 and variants within 400kb on chromosome 6. D. Crystal structure of HCRTR2 (PDB ID 6TPJ) showing localization of rs2653349 that changes Isoleucine to Phenylalanine or to valine at the transmembrane domain of HCRTR2. Protein sequence was visualized using iCn3D (https://www.ncbi.nlm.nih.gov/Structure/icn3d/full.html). The variant rs2653349 was aligned with the sequence (arrows to Human Missense variant in Figure) and the previously published canine HCRTR2 mutations^96^, which disrupt transmembrane and signaling domains or truncate the HCRTR2 protein are highlighted in cyan. E. Forest plot of associations between the A allele of genetic variant rs2653349 in *HCRTR2* and the colocalized sleep phenotypes.

Finally, multi-trait clustering suggested the possibility of at least three heterogeneous pathways influencing daytime napping. Bayesian nonnegative matrix factorization (bNMF)^39^ clustering for 123 variants with 17 self-reported and accelerometer-derived sleep traits identified 3 clusters (63% of 1,000 iterations) and these same 3 clusters were also present in an additional 34% of iterations with 4 clusters (**Table 1, Supplementary Table 14, Supplementary Figure 3a**) reflecting a) “sleep propensity” (cluster 1; 6 contributing loci with *CRHR1, SKOR2, KSR2, ASCL4, RERE*, and *ECE2*); b) “disrupted sleep” (cluster 2; 5 contributing loci with *SHISA4, ADO, NRXN3, FNDC5*, and *GS1-259H13*.*13* as lead); and c) “early sleep timing” (cluster 3; 9 contributing loci with *HCRTR2, ALG10, ALG10B, PATJ, BTBD9, MTNR1B, AGAP1, RP11-6N13*.*1*, and *ZBTB5* as lead loci, notably not at known core clock genes).

**Table 1.**
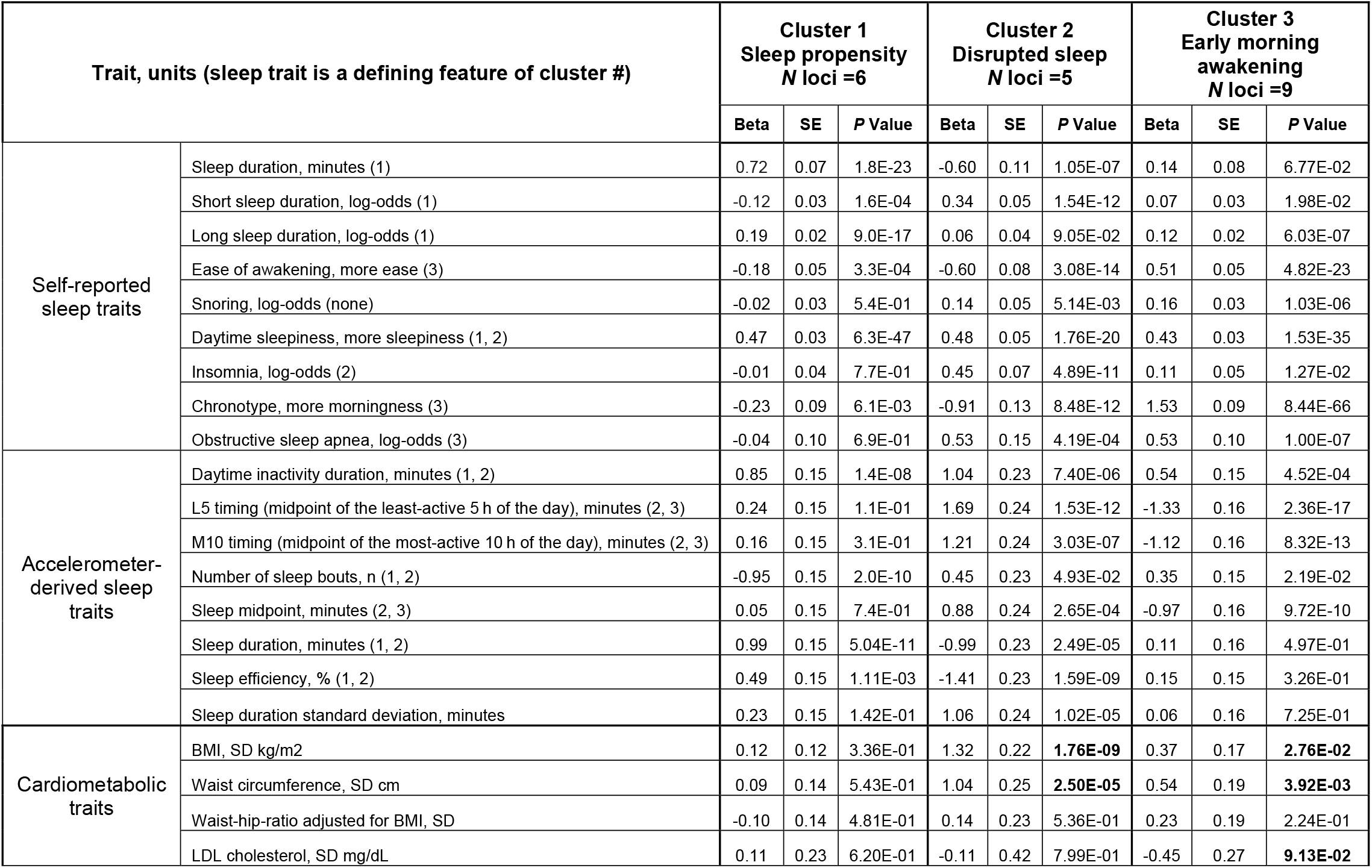
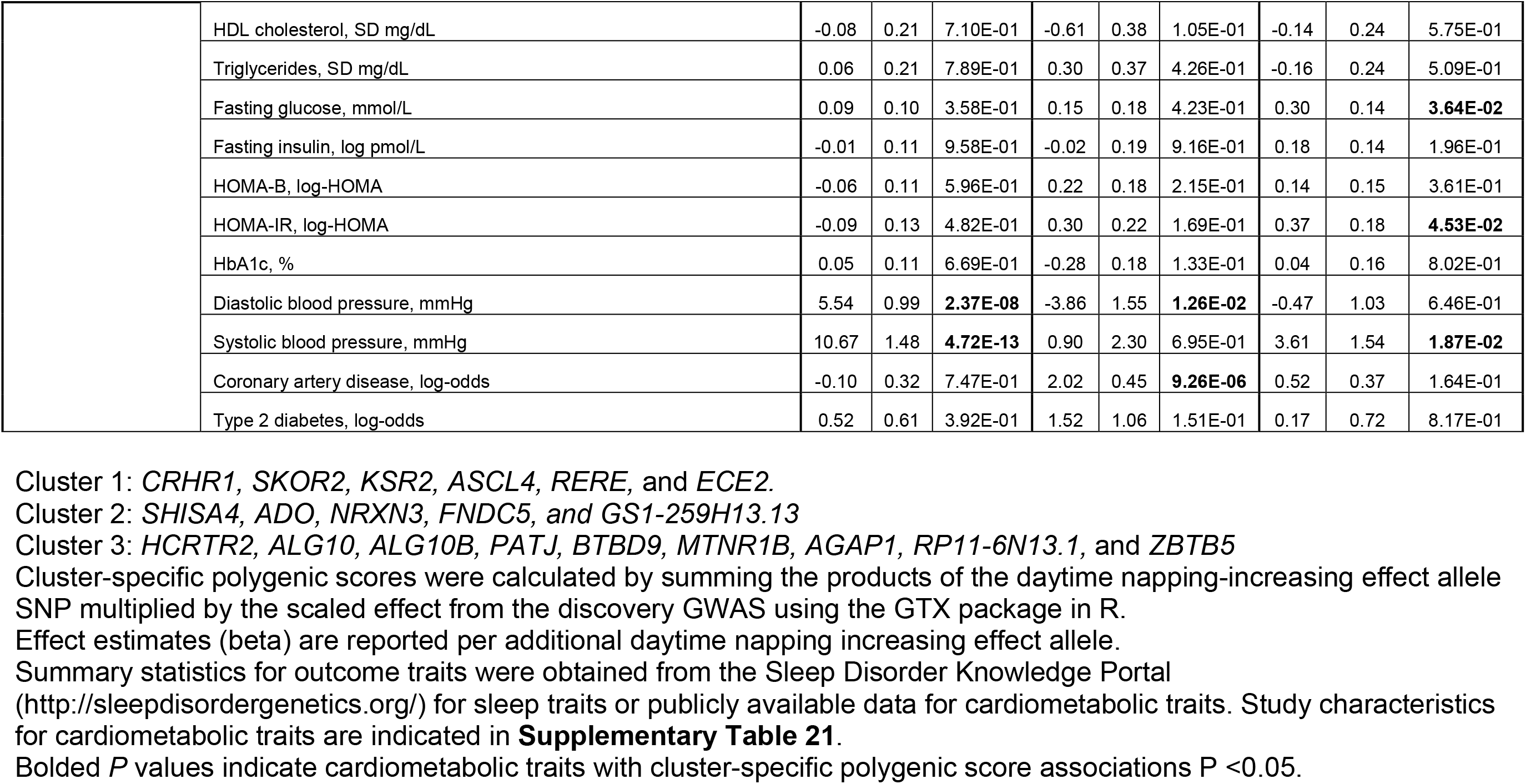
Cluster-specific daytime napping polygenic scores associations with self-reported and accelerometer-derived sleep traits and other cardiometabolic traits.

| Trait, units (sleep trait is a defining feature of cluster #) |  | Cluster 1<br>Sleep propensity<br>N loci =6 |  |  | Cluster 2<br>Disrupted sleep<br>N loci =5 |  |  | Cluster 3<br>Early morning<br>awakening<br>N loci =9 |  |  |
| --- | --- | --- | --- | --- | --- | --- | --- | --- | --- | --- |
|  |  | Beta | SE | P Value | Beta | SE | P Value | Beta | SE | P Value |
| Self-reported<br>sleep traits | Sleep duration, minutes (1) | 0.72 | 0.07 | 1.8E-23 | -0.60 | 0.11 | 1.05E-07 | 0.14 | 0.08 | 6.77E-02 |
|  | Short sleep duration, log-odds (1) | -0.12 | 0.03 | 1.6E-04 | 0.34 | 0.05 | 1.54E-12 | 0.07 | 0.03 | 1.98E-02 |
|  | Long sleep duration, log-odds (1) | 0.19 | 0.02 | 9.0E-17 | 0.06 | 0.04 | 9.05E-02 | 0.12 | 0.02 | 6.03E-07 |
|  | Ease of awakening, more ease (3) | -0.18 | 0.05 | 3.3E-04 | -0.60 | 0.08 | 3.08E-14 | 0.51 | 0.05 | 4.82E-23 |
|  | Snoring, log-odds (none) | -0.02 | 0.03 | 5.4E-01 | 0.14 | 0.05 | 5.14E-03 | 0.16 | 0.03 | 1.03E-06 |
|  | Daytime sleepiness, more sleepiness (1, 2) | 0.47 | 0.03 | 6.3E-47 | 0.48 | 0.05 | 1.76E-20 | 0.43 | 0.03 | 1.53E-35 |
|  | Insomnia, log-odds (2) | -0.01 | 0.04 | 7.7E-01 | 0.45 | 0.07 | 4.89E-11 | 0.11 | 0.05 | 1.27E-02 |
|  | Chronotype, more morningness (3) | -0.23 | 0.09 | 6.1E-03 | -0.91 | 0.13 | 8.48E-12 | 1.53 | 0.09 | 8.44E-66 |
|  | Obstructive sleep apnea, log-odds (3) | -0.04 | 0.10 | 6.9E-01 | 0.53 | 0.15 | 4.19E-04 | 0.53 | 0.10 | 1.00E-07 |
| Accelerometer-derived sleep<br>traits | Daytime inactivity duration, minutes (1, 2) | 0.85 | 0.15 | 1.4E-08 | 1.04 | 0.23 | 7.40E-06 | 0.54 | 0.15 | 4.52E-04 |
|  | L5 timing (midpoint of the least-active 5 h of the day), minutes (2, 3) | 0.24 | 0.15 | 1.1E-01 | 1.69 | 0.24 | 1.53E-12 | -1.33 | 0.16 | 2.36E-17 |
|  | M10 timing (midpoint of the most-active 10 h of the day), minutes (2, 3) | 0.16 | 0.15 | 3.1E-01 | 1.21 | 0.24 | 3.03E-07 | -1.12 | 0.16 | 8.32E-13 |
|  | Number of sleep bouts, n (1, 2) | -0.95 | 0.15 | 2.0E-10 | 0.45 | 0.23 | 4.93E-02 | 0.35 | 0.15 | 2.19E-02 |
|  | Sleep midpoint, minutes (2, 3) | 0.05 | 0.15 | 7.4E-01 | 0.88 | 0.24 | 2.65E-04 | -0.97 | 0.16 | 9.72E-10 |
|  | Sleep duration, minutes (1, 2) | 0.99 | 0.15 | 5.04E-11 | -0.99 | 0.23 | 2.49E-05 | 0.11 | 0.16 | 4.97E-01 |
|  | Sleep efficiency, % (1, 2) | 0.49 | 0.15 | 1.11E-03 | -1.41 | 0.23 | 1.59E-09 | 0.15 | 0.15 | 3.26E-01 |
|  | Sleep duration standard deviation, minutes | 0.23 | 0.15 | 1.42E-01 | 1.06 | 0.24 | 1.02E-05 | 0.06 | 0.16 | 7.25E-01 |
| Cardiometabolic<br>traits | BMI, SD kg/m2 | 0.12 | 0.12 | 3.36E-01 | 1.32 | 0.22 | **1.76E-09** | 0.37 | 0.17 | **2.76E-02** |
|  | Waist circumference, SD cm | 0.09 | 0.14 | 5.43E-01 | 1.04 | 0.25 | **2.50E-05** | 0.54 | 0.19 | **3.92E-03** |
|  | Waist-hip-ratio adjusted for BMI, SD | -0.10 | 0.14 | 4.81E-01 | 0.14 | 0.23 | 5.36E-01 | 0.23 | 0.19 | 2.24E-01 |
|  | LDL cholesterol, SD mg/dL | 0.11 | 0.23 | 6.20E-01 | -0.11 | 0.42 | 7.99E-01 | -0.45 | 0.27 | **9.13E-02** |
|  | HDL cholesterol, SD mg/dL | -0.08 | 0.21 | 7.10E-01 | -0.61 | 0.38 | 1.05E-01 | -0.14 | 0.24 | 5.75E-01 |
|  | Triglycerides, SD mg/dL | 0.06 | 0.21 | 7.89E-01 | 0.30 | 0.37 | 4.26E-01 | -0.16 | 0.24 | 5.09E-01 |
|  | Fasting glucose, mmol/L | 0.09 | 0.10 | 3.58E-01 | 0.15 | 0.18 | 4.23E-01 | 0.30 | 0.14 | **3.64E-02** |
|  | Fasting insulin, log pmol/L | -0.01 | 0.11 | 9.58E-01 | -0.02 | 0.19 | 9.16E-01 | 0.18 | 0.14 | 1.96E-01 |
|  | HOMA-B, log-HOMA | -0.06 | 0.11 | 5.96E-01 | 0.22 | 0.18 | 2.15E-01 | 0.14 | 0.15 | 3.61E-01 |
|  | HOMA-IR, log-HOMA | -0.09 | 0.13 | 4.82E-01 | 0.30 | 0.22 | 1.69E-01 | 0.37 | 0.18 | **4.53E-02** |
|  | HbA1c, % | 0.05 | 0.11 | 6.69E-01 | -0.28 | 0.18 | 1.33E-01 | 0.04 | 0.16 | 8.02E-01 |
|  | Diastolic blood pressure, mmHg | 5.54 | 0.99 | **2.37E-08** | -3.86 | 1.55 | **1.26E-02** | -0.47 | 1.03 | 6.46E-01 |
|  | Systolic blood pressure, mmHg | 10.67 | 1.48 | **4.72E-13** | 0.90 | 2.30 | 6.95E-01 | 3.61 | 1.54 | **1.87E-02** |
|  | Coronary artery disease, log-odds | -0.10 | 0.32 | 7.47E-01 | 2.02 | 0.45 | **9.26E-06** | 0.52 | 0.37 | 1.64E-01 |
|  | Type 2 diabetes, log-odds | 0.52 | 0.61 | 3.92E-01 | 1.52 | 1.06 | 1.51E-01 | 0.17 | 0.72 | 8.17E-01 |

Cluster 1: *CRHR1*, *SKOR2*, *KSR2*, *ASCL4*, *RERE*, and *ECE2*.
Cluster 2: *SHISA4*, *ADO*, *NRXN3*, *FNDC5*, and *GS1-259H13.13*
Cluster 3: *HCRTR2*, *ALG10*, *ALG10B*, *PATJ*, *BTBD9*, *MTNR1B*, *AGAP1*, *RP11-6N13.1*, and *ZBTB5*
Cluster-specific polygenic scores were calculated by summing the products of the daytime napping-increasing effect allele SNP multiplied by the scaled effect from the discovery GWAS using the GTX package in R.
Effect estimates (beta) are reported per additional daytime napping increasing effect allele.
Summary statistics for outcome traits were obtained from the Sleep Disorder Knowledge Portal
Bolded *P* values indicate cardiometabolic traits with cluster-specific polygenic score associations $P < 0.05$ .

A fourth possible cluster “obstructive sleep apnea” was observed in 34% of 1,000 iterations (**Supplementary Figure 3b**). Results agreed with the dendrogram of an alternative unsupervised hierarchical clustering method^23^ (**Supplementary Figure 3c**), and clusters 1 and 2 partly overlapped with previously observed clusters for daytime sleepiness^23^.

### Genes at association signals are enriched in brain and GABAergic neurons, and in neural development and opioid signaling pathways

In order to identify tissues, neuronal subtypes and annotated pathways relevant to daytime napping, we first mapped the genes near association signals and then tested for their over-representation relative to all genes in experimental genome-wide datasets. Gene-based associations for 21,761 genes mapped with Pascal^40^ are listed in **Supplementary Table 15**; 324 genes showed association after Bonferroni correction. The identified signals were enriched for genes predominantly expressed in brain tissues, including the frontal cortex (*P* =1.18×10^−7^ and nucleus accumbens (*P* =1.26×10^−7^) (**Figure 3a, Supplementary Table 16**). Single cell enrichment analyses in FUMA^41^ using human brain datasets (listed in **Figure 3b**) showed consistent enrichment in GABAergic neurons across several brain tissues including the prefrontal cortex and midbrain. In addition, pathway enrichment analysis using MAGMA^42^ and Pascal^40^ indicated enrichment of genes involved in regulation of transmission across chemical synapses, neuronal system, opioid signaling, among others (**Figure 3c, Supplementary Tables 17,18**).

**Figure 3.**
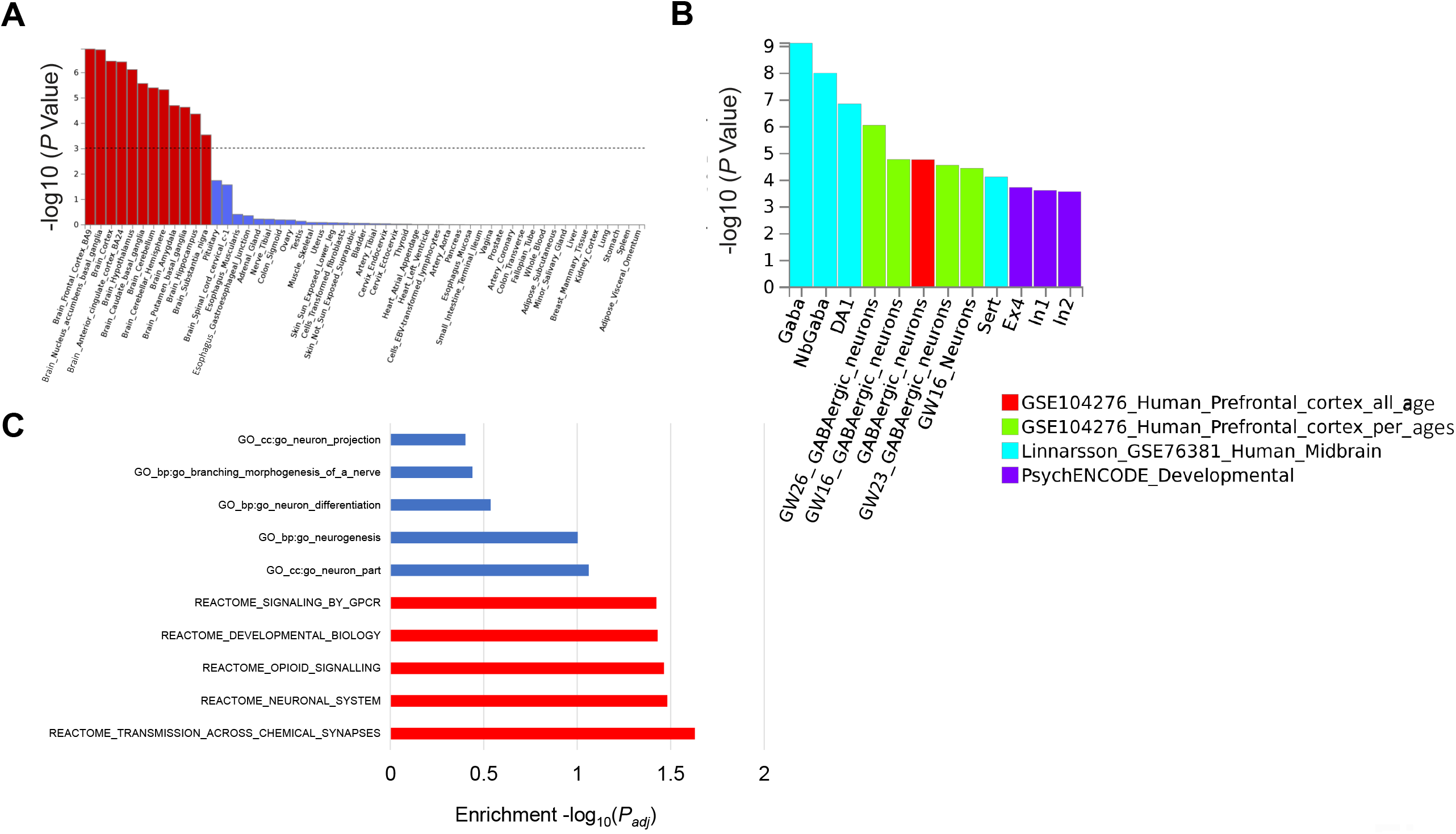
Tissue expression, single-cell, and pathway-based enrichment analyses for daytime napping. A. MAGMA tissue expression analysis using gene expression per tissue based on GTEx RNA-seq data for 53 specific tissue types. Significant tissues are shown in red (*P* <9.43×10^−4^). B. Significant single cell types from single cell enrichment analyses using human brain datasets in FUMA. C. Top 5 pathways determined from analysis using MAGMA gene sets and Pascal (gene-set enrichment analysis using 1077 pathways from KEGG, REACTOME, BIOCARTA databases). Significant pathways are shown in red. All pathway and tissue expression analyses in this figure can be found in tabular form in Supplementary Tables 16, 17, 18.

### The genetic contributors to daytime napping are shared with cardiometabolic diseases and behavior traits

To gain insights into shared heritability of daytime napping with other disease and behavior traits, we performed cross-trait LD score regression^26^ using publicly available GWAS data for 257 traits. Modest positive correlations were observed between daytime napping and several anthropometric, cardiometabolic, and psychiatric disease traits including BMI, type 2 diabetes, and depressive symptoms (**Figure 4a, Supplementary Table 19**). To further characterize shared genetic links between daytime napping and diseases in a disease-enriched and independent health system-based clinical cohort, we conducted a phenome-wide association study (PheWAS) in the Partners Biobank (*n* =23,561 participants of European ancestry with genetic data)^43,44^. We generated a daytime napping genome-wide polygenic score (GPS) and tested associations with 951 ICD-code based disease categories. PheWAS showed 3 Bonferroni-significant associations (18 FDR-significant), including positive associations with essential hypertension (phecode: 401.1; GPS q10 vs q1 odds ratio [95% confidence interval]: 1.30 [1.13, 1.51]), obesity (phecode: 278.1; GPS q10 vs q1: 1.38 [1.18, 1.62]), and chronic nonalcoholic liver disease (phecode: 571.5; GPS q10 vs q1: 1.51 [1.18, 1.92]), which encompasses diagnosis codes for chronic non-specific or non-alcoholic liver disease (**Figure 4b,c, Supplementary Table 20**). In support, we also observed associations of a polygenic score of the 123 napping variants, and polygenic sub-scores for each of the 3 clusters with cardiometabolic traits from large-scale public GWAS, (**Table 1, Supplementary Table 21**). Cluster-specific polygenic score associations varied across outcomes, and included associations with higher blood pressure for cluster 1, and adiposity traits for clusters 2 and 3 (**Table 1**)

**Figure 4.**
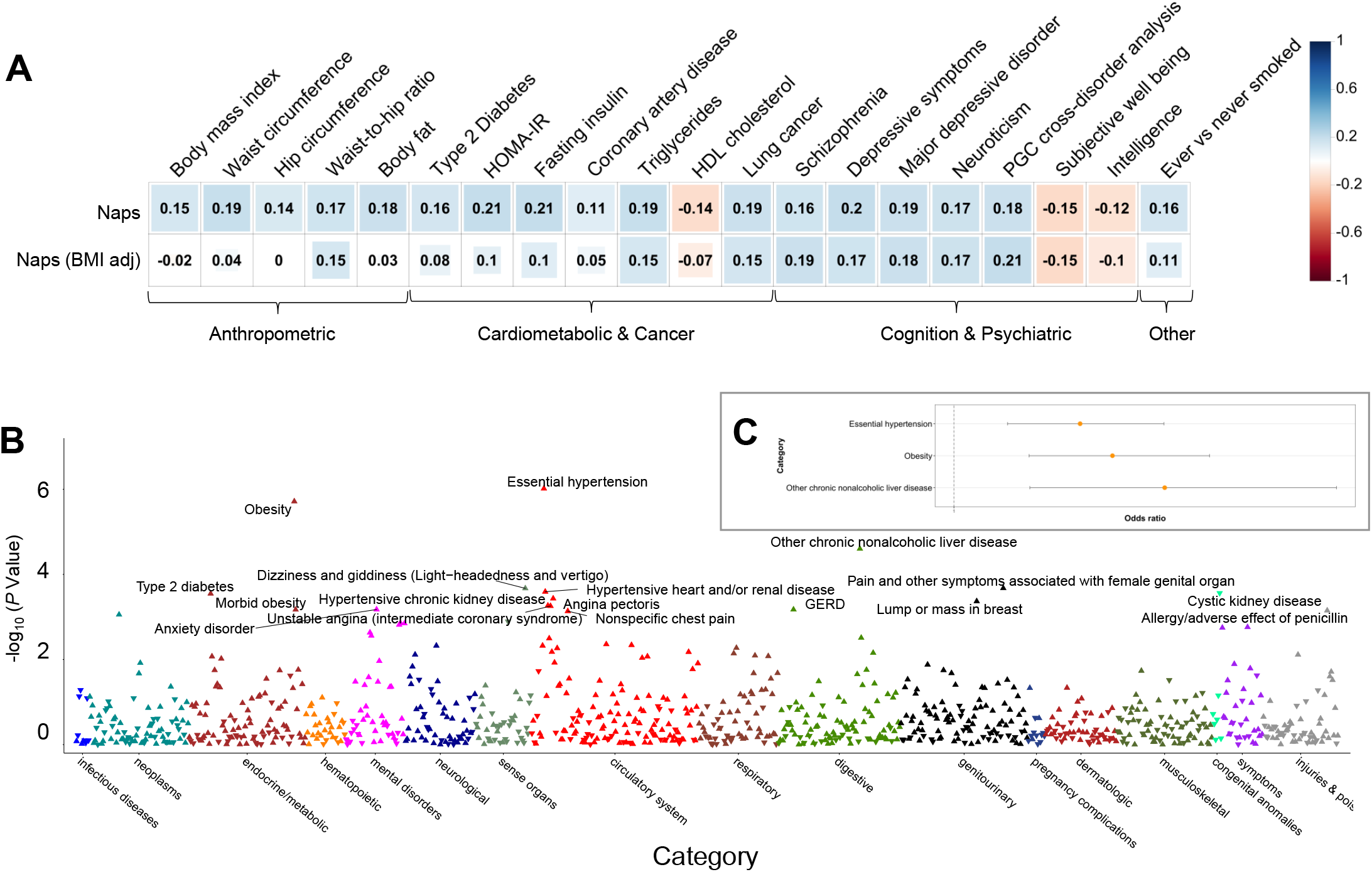
Genome-wide genetic architecture of daytime napping correlations and associations with diseases and behavioral traits. A. Shared genetic architecture between daytime napping and diseases and selected behavioral traits. Linkage disequilibrium (LD) score regression estimates of genetic correlation (*r*_g_) were obtained by comparing genome-wide association estimates for daytime napping with summary statistics estimates from 257 publicly available genome-wide association studies. Blue indicates positive genetic correlation and red indicates negative genetic correlation; *r*_g_ values are displayed for significant correlations. Larger colored squares correspond to more significant *P* values. Full genetic correlations for all 257 traits can be found in Supplementary Table 19. B. Manhattan plot of phenome-wide association findings for daytime napping genome-wide polygenic score in Partners Biobank (*n* =23,561). The x-axis is color-coded phecodes organized by broad disease categories and the y-axis is *P* value of association (-log_10_ *P*). The horizontal red line depicts phenome-wide significance using Bonferroni correction for all tested diseases (951 diseases), and the horizontal blue line depicts phenome-wide significance using False Discovery Rate (FDR) correction. Upward arrows denote positive associations (OR >1), and downward arrows denote negative associations (OR <1). Full results for all 951 diseases can be found in Supplementary Table 20. C. Cross-sectional association between quartile 10 and quartile 1 of daytime napping genome-wide polygenic score and essential hypertension, obesity, and chronic nonalcoholic liver disease in the Partners Biobank (*n* =23,561). Error bars represent the 95% confidence intervals for association.

### Mendelian randomization suggests a causal effect of increased napping frequency on increased blood pressure and waist circumference

To address the question of whether daytime napping is causal for cardiometabolic diseases, we performed two-sample Mendelian randomization (MR) analyses using the 123 loci as unconfounded genetic instruments with 15 cardiometabolic traits (**Supplementary Table 21**). A causal effect of increased daytime napping frequency on a range of adverse cardiometabolic outcomes was observed (**Figure 5a,b**), including higher diastolic blood pressure (DBP; 0.25 standard deviation (SD) increase in mmHg per category increase in daytime napping, 95% CI [0.15, 0.34], *P* =2.99×10^−7^), systolic blood pressure (SBP; 0.18 SD, [0.09, 0.27], *P* =5.15×10^−5^), and waist circumference (0.28 SD units, [0.11, 0.45], *P* =1.3×10^−3^), all of which surpassed multiple testing correction. As these effects may be explained by pleiotropic effects of these variants on pathways independent of napping, we conducted four sensitivity analyses robust to some forms of unbalanced horizontal pleiotropy and found consistent effects (**Supplementary Table 22-23; Supplementary Figure 4**).

**Figure 5.**
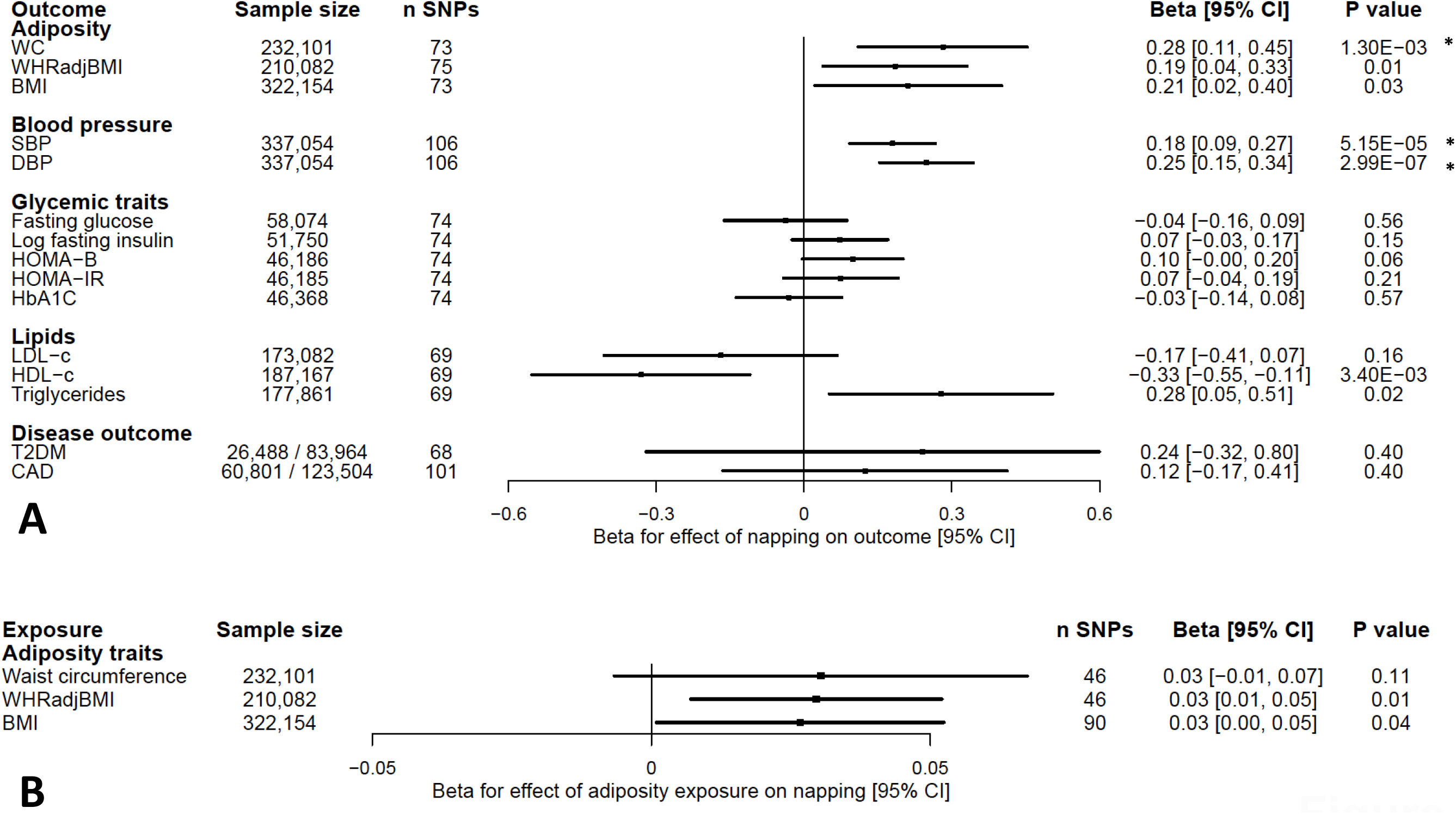
Mendelian randomization reveals a causal effect of increased genetic liability to daytime napping on greater waist circumference and blood pressure. *: significant at Bonferroni-corrected alpha threshold and robust in sensitivity analyses BMI: body mass index; CAD: coronary artery disease; CI: confidence interval; DBP: diastolic blood pressure; HOMA: Homeostatic Model Assessment of Insulin Resistance; HOMAB: homeostasis model assessment of β-cell function; LDL: low density lipoprotein; HDL: high density lipoprotein OR: odds ratio; SBP: systolic blood pressure; SNP: single nucleotide polymorphism; T2DM: type 2 diabetes mellitus; WC: waist circumference; WHRadjBMI: waist-to-hip ratio adjusted for BMI. The effect of the genetic instrument on each outcome was calculated using random-effects inverse-variance weighted meta-analysis. Daytime napping exposure represents a one-unit increase in napping category (never, sometimes, usually). Black boxes show effect estimates, and surrounding lines display 95% confidence intervals. (A) IVW effect estimates for increased daytime napping on cardiometabolic outcomes and risk factors (note that a unit increase in the adiposity and blood pressure measurement represents a standard deviation increase), and (B) IVW effect estimates for adiposity traits on daytime napping.

Given prior evidence for a causal effect of higher BMI on daytime sleepiness^23^, we tested the hypothesis that adiposity traits [waist circumference, waist-to-hip ratio adjusted for BMI (WHRadjBMI), and BMI] influenced daytime napping propensity. Only genetically instrumented WHRadjBMI was nominally associated with a modest increase in daytime napping (inverse variance weighted: 0.03 category increase in daytime napping per SD increase in waist-to-hip ratio, [0.01, 0.05], *P* =0.01) (**Figure 5b, Supplementary Table 22**).

### Leveraging HCRTR1 and HCRTR2 genetic associations to assess the cardiovascular safety profile of dual orexin antagonists

Given our observation that the hypocretin arousal pathway was involved in daytime napping propensity (genetic variants at *HCRTR1* and *HCRTR2*), recent reports suggesting that mammalian orexin signaling has cardioprotective effects^45^, and the pharmacologic use of dual orexin receptor antagonists (DORAs) as sleep medications, we wondered whether our variants may serve as genetic instruments to test the cardiovascular safety of orexin receptors as drug targets. To test for such potential on-target cardiovascular side effects, we used missense variants in *HCRTR1* and *HCRTR2* (A allele of rs2271933 (*HCRTR1*); A allele of rs2653349 (rs2653349)), both associated with more frequent daytime napping and daytime sleepiness^23^, as proxies for pharmacologic inhibition of these proteins, and tested for associations with cardiovascular phenotypes in large GWAS **(Supplementary Table 24)**. This analysis revealed no associations with cardiovascular outcomes (**Figure 6**), and opposing effects on systolic blood pressure at *HCRTR1* (Beta = -0.10 mmHg, 95% CI [-0.17, - 0.04], *P* =1.00×10^−4^) and *HCRTR2* (Beta = 0.14 mmHg, 95% CI [0.07, 0.21], *P* =1.00×10^−04^). We supplemented this analysis with a hypothesis-free scan across 1,402 ICD-code defined phenotypes in the UK Biobank^46^ and found no variant-disease associations (**Supplementary Figure 5; Supplementary Table 25**). Human genetic evidence therefore does not support a net excess adverse cardiovascular risk from on-target inhibition of *HCRTR1* and *HCRTR2*. These results may also inform on-target side-effects for orexin receptor agonists currently in development for narcolepsy.

**Figure 6.**
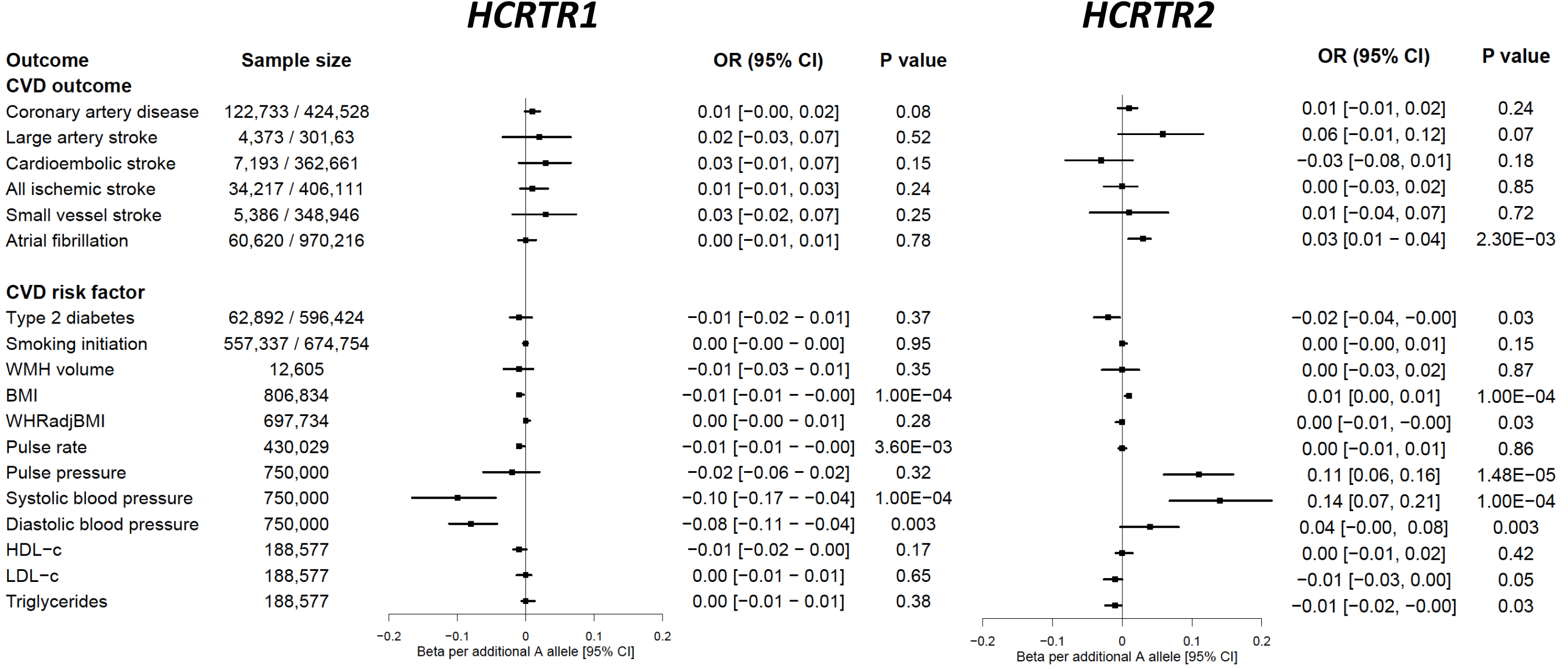
Pharmacogenetic phenome-wide association study for missense variants in *HCRTR1* and *HCRTR2*, which encode targets of Suvorexant, an FDA-approved sleep medication with an unknown cardiovascular safety profile. Additional details regarding the included studies are provided in Supplementary Tables 25-26.

## Discussion

We comprehensively investigated the genomic influences of daytime napping using the largest discovery and replication sample sizes to date. We identified 123 independent loci in the UK Biobank with strong evidence of replication in 23andMe, an independent study with different demographic characteristics. Variant effects were largely independent of BMI, and the associations retained significance when GWAS was restricted to healthier participants, suggesting that signals were not driven by poor health. Our results advance the biology of daytime napping, refine the understanding of pleiotropy and causality in the relationship with sleep and cardiometabolic traits, and inform pharmacologic investigations of orexin antagonism.

Our genomic findings indicate that daytime napping is a distinct sleep phenotype with both unique biological determinants (26/123 loci) as well as biological determinants shared with other sleep traits, primarily daytime sleepiness and sleep duration. The identified variants highlight a central role for arousal-regulating neuropeptide signaling pathways in daytime napping propensity. Most prominent among these pathways was the well-established hypocretin arousal pathway^47^ (including missense variants in *HCRTR1, HCRTR2*, and *TRPC6*). It is thus possible that orexin receptor agonists, currently under development for narcolepsy, may have roles for treatment of the continuum of impaired arousal. Additional pathways with known roles in sleep/wake biology in model organisms^48^ include neuronal excitability driven by variation in the function of potassium channels and glutamate signaling, EGFR signaling pathway, and opioid signaling. Notably, expression of genes under association peaks was most enriched in the frontal cortex, similar to observations for daytime inactivity duration^19^, and other brain regions prominently implicated in sleep duration, timing and quality traits^22,24,25^. While functional studies are needed to understand how genetic variants at each locus influence daytime napping, cross-trait clustering of the identified loci suggest at least three underlying physiologic mechanisms, including 1) propensity for longer sleep, 2) consequence of poor and disrupted sleep, and 3) napping concomitant with early sleep timing, potentially reflecting loss of function in arousal pathways. Notably, genetic links between daytime and sleep disorders, e.g. sleep apnea or restless legs syndrome, may be partially undetected by our study because of poor assessment of these disorders in the UK Biobank.

We found that the genetic architecture of daytime napping is shared with cardiometabolic traits and diseases, consistent with previous epidemiologic associations of frequent daytime napping with cardiometabolic risk^7–13,49^. At the locus level, we observed colocalization of the daytime napping loci with daytime sleepiness, snoring, chronotype, and BMI loci at *PNOC* and *PATJ*, suggesting an obesity-hypersomnolence pathway^50^. Furthermore, colocalization of *FADS1* gene expression in the frontal cortex with the daytime napping signal, and genetic correlations with HDL cholesterol and lipid subfractions suggest uncharacterized pleiotropic roles of lipid metabolism on sleep. Positive genome-wide genetic correlations were observed with multiple anthropometric, glycemic, and cardiometabolic traits. In a large health system-based clinical cohort, phenome-wide association analyses using a daytime napping genome-wide polygenic score further supported associations with obesity and hypertension, in addition to other cardiometabolic diseases.

A key question is whether habitual daytime napping has detrimental or beneficial chronic effects on cardiometabolic health, and therefore we leveraged two-sample Mendelian randomization to explore causal relationships. Our main findings suggest potentially deleterious effects of daytime napping on cardiometabolic health, with prominent effects on increased blood pressure and waist circumference. A causal link with higher blood pressure is in agreement with earlier epidemiologic findings between self-reported and actigraphy-measured daytime napping and hypertension^51–53^.

Mechanisms driving this relationship are unknown but may include chronic effects of short and/or poor quality sleep linked to napping, or chronic effects related to transient evening blood pressure surges following daytime napping, as is observed with morning awakening surges, a risk factor for cardiovascular events^54,55^. Similarly, mechanisms underlying the link between daytime napping and visceral adiposity are poorly understood^56^. Although results from the MR Egger sensitivity analysis of waist circumference on napping were inconsistent with findings from our primary MR analysis, the genetic overlap we demonstrate with BMI indicates that the Egger analysis may be biased by violation of the “instrument strength independent of direct effect (INSIDE)” assumption^57^. Polygenic scores of each napping subtype showed heterogeneous associations with cardiometabolic outcomes across clusters, including associations with higher systolic and diastolic blood pressure for cluster 1 (also associated with longer sleep duration), and other adiposity traits for the remaining clusters. Exploring causal relationships with biologically distinct subtypes of daytime napping will be important to understand the beneficial or detrimental role of different aspects of napping biology with disease outcomes. As yet, not enough genetic factors for biologically distinct subtypes of naps have been identified to robustly test these relationships.

Because our study identified naps-inducing (likely partial loss of function) variants in both orexin receptors that have emerged as novel drug targets for insomnia, and because prior work has linked the orexin system to cardiovascular health^45^, we further leveraged this coding variation to predict the cardiovascular consequences of long-term pharmacologic modulation of HCRTR1 and HCRTR2. We found no net effect of inhibition of orexin receptors on cardiovascular outcomes, nor on any ICD-code defined disease outcomes in PheWAS, supporting the safety of these orexin antagonist therapies. A novel association of *HCRTR1* and *HCRTR2* with blood pressure was observed, however the direction of effect relative to the daytime napping effect differed for the two variants. This suggests a neutral net blood pressure effect of DORAs, and more broadly suggests pleiotropic effects of these proteins on blood pressure regulation. However, it is possible that these genetic variants do not proxy peripheral effects (e.g. bone marrow) of Suvorexant on HCRTR1 and HCRTR2 inhibition^45^. Notably, this is the first application of PheWAS to study on-target side effects of sleep medications, and sets the stage for future advances in the understanding of the health consequences of orexin antagonism.

Our analyses are limited by the crude assessment of daytime napping frequency via questionnaire with no information on duration or timing. Replication of most loci and the specific association with daytime activity duration as measured by accelerometer, however, support our findings. It remains possible that rare and structural variation have a strong contribution to the genetic architecture of daytime napping, however these data were not tested in the present analysis. Finally, despite consistency in Mendelian randomization estimates, these analyses require strong, unverifiable assumptions for the determination of causality and therefore require confirmation in randomized controlled trials of sleep interventions. In summary, our genetic analyses contribute important insight into the biology and cardiometabolic consequence of habitual daytime napping in adults.

## Methods

### UK Biobank

The UK Biobank is a large population-based study established to allow detailed investigations of the genetic and lifestyle determinants of a wide range of phenotypes^58^. Data from >500,000 participants living in the United Kingdom who were aged 40-69 and living <25 miles from a study center participated in the study between 2006 and 2010. Extensive phenotypic data were self-reported upon baseline assessment by participants using touchscreen tests and questionnaires and at nurse-led interviews. The UK Biobank study was approved by the National Health Service National Research Ethics Service (ref. 11/NW/0382), and all participants provided written informed consent to participate. The current study was conducted under UK Biobank application 6818.

### Daytime napping, covariates, and other self-reported and objectively measured sleep traits

At baseline assessment, all study participants self-reported daytime napping (*n* =501,646). Participants were asked *Do you have a nap during the day?*, with responses *Never/rarely, Sometimes, Usually, Prefer not to answer*. Responses were treated as a continuous variable in the GWAS. *Prefer not to answer* responses were set to missing.

Participants further self-reported age, gender, sleep duration, chronotype, insomnia symptoms, sleep apnea, smoking, and overall health. Weight, height, and waist circumference were measured and body-mass index (BMI) was calculated as weight (kg) / height^2^(m^2^). Systolic and diastolic blood pressure were measured at baseline and the average of two automated readings was used. Socio-economic status was represented by the Townsend deprivation index based on national census data immediately preceding participation in the UK Biobank. Assessment season was determined from baseline assessment visit date and categorized as 1 for winter [January to March], 2 for spring [April to June], 3 for summer [July to September], and 4 for fall [October to December], as previously conducted^59^. Participants rated their overall health in response to the question, *In general how would you rate your overall health?*, with responses *excellent, good, fair, poor, do not know*, and *prefer not to answer*. Cases of sleep apnea were determined from self-report during nurse-led interviews or health records using International Classification of Diseases (ICD)-10 codes for sleep apnea (G47.3). Insomnia symptoms were ascertained from self-report to the question, *Do you have trouble falling asleep at night or do you wake up in the middle of the night?* with responses never/rarely, sometimes, usually, prefer not to answer. Participants who responded *usually* were set as insomnia cases, and remaining participants were set as controls. Smoking status (*never, former, current*) was further self-reported. Missing covariates were imputed by using sex-specific median values for continuous variables (*i*.*e*., BMI and Townsend index).

A subset of 103,711 participants from the UK Biobank wore actigraphy devices (Axivity AX3) for up to 7 days, approximately 2.8 to 9.7 years after their study baseline visits. Details on quality control and data processing have been described previously^19,60^. The following sleep measures were derived by processing raw accelerometer data: daytime inactivity duration, sleep duration, sleep efficiency, number of sleep bouts within the sleep period time window, sleep midpoint, midpoint of the least-active 5 h of the day (L5 timing) and midpoint of the most-active 10 h of the day (M10 timing). Specifically, daytime inactivity duration was estimated by the total daily duration of estimated bouts of inactivity that fell outside of the sleep period time window. These inactivity bouts are any inactivity lasting ≥30 minutes. Inactivity bouts that are <60 minutes apart are combined to form inactivity blocks. As previously described, this measure captures very inactive states such as napping and wakeful rest but not inactivity such as sitting and reading or watching television, which are associated with a low but detectable level of movement^19^.

### Genome-wide association study for daytime napping in UK Biobank

Genotyping was performed by the UK Biobank, and genotyping, quality control, and imputation procedures are described in detail previously^61^. In brief, blood, saliva, and urine were collected from participants, and DNA was extracted from the buffy coat samples. Participant DNA was genotyped on two arrays, UK BiLEVE and UK Biobank Axiom with >95% common content and genotypes for ∼800,000 autosomal SNPs were imputed to two reference panels. Genotypes were called using Affymetrix Power Tools software. Sample and SNPs for quality control were selected from a set of 489,212 samples across 812,428 unique markers. Sample quality control (QC) was conducted using 605,876 high quality autosomal markers. Samples were removed for high missingness or heterozygosity (968 samples) and sex chromosome abnormalities (652 samples). Genotypes for 488,377 samples passed sample QC (∼99.9% of total samples). Marker based QC measures were tested in the European ancestry subset (n=463,844), which was identified based on principal components of ancestry. SNPs were tested for batch effects (197 SNPs/batch), plate effects (284 SNPs/batch), Hardy-Weinberg equilibrium (572 SNPs/batch), sex effects (45 SNPs/batch), array effects (5417 SNPs), and discordance across control replicates (622 on UK BiLEVE Axiom array and 632 UK Biobank Axiom array; p-value <10^−12^ or <95% for all tests). For each batch (106 batches total) markers that failed at least one test were set to missing. Before imputation, 805,426 SNPs pass QC in at least one batch (>99% of the array content).

Population structure was captured by principal component analysis on the samples using a subset of high quality (missingness <1.5%), high frequency SNPs (>2.5%) (∼100,000 SNPs) and identified the sub-sample of white British descent. In addition to the calculated population structure by the UK Biobank, we locally further clustered subjects into four ancestry clusters using K-means clustering on the principal components, identifying 453,964 subjects of European ancestry. For the current analysis, individuals of non-white ethnicity were excluded to limit confounding effects. The UK Biobank centrally imputed autosomal SNPs to UK10K haplotype, 1000 Genomes Phase 3, and Haplotype Reference Consortium (HRC). Autosomal SNPs were pre-phased using SHAPEIT3 and imputed using IMPUTE4. In total ∼96 million SNPs were imputed. Related individuals were identified by estimating kinship coefficients for all pairs of samples, using only markers weakly informative of ancestral background.

Genetic association analysis for daytime napping (*never/rarely, sometimes*, and *always*) was performed in related subjects of European ancestry with self-reported daytime napping data (*n* =452,633) using BOLT-LMM^62^ linear mixed models and an additive genetic model adjusted for age, sex, 10 principal components of ancestry, genotyping array and genetic correlation matrix [jl2] with a maximum *per* SNP missingness of 10% and *per* sample missingness of 40%. We used a hard-call genotype threshold of 0.1, SNP imputation quality threshold of 0.80, and a MAF threshold of 0.001, and 13,304,133 genetic variants remained.

Trait heritability was calculated as the proportion of trait variance due to additive genetic factors measured in this study using BOLT-REML^62^, to leverage the power of raw genotype data together with low frequency variants (MAF ≥0.001). Lambda inflation (λ) values were calculated using GenABEL in R, and estimated values were consistent with those estimated for other highly polygenic complex traits. Furthermore, follow-up GWAS for daytime napping were conducted using BOLT-LMM^62^ and included sensitivity analyses restricted to participants self-reporting *excellent* or *good* overall health (*n* =338,764), GWAS adjusting for BMI in addition to baseline adjustments, and lastly sex-stratified GWAS (male *n* =207,108; female *n* =245,525).

Distinct genomic risk *loci* were defined using FUMA v1.3.3 on basis of genome-wide significance (*P* <5×10^−8^) and whether independent from each other (*r*^2^ < 0.6) within a 1 Mb window. Annotation of the lead variants, including predicted sequence consequence, was obtained from the FUMA output. We also determined the PICS probability for each lead variant being the causal variant at the locus^63^.

For the 123 lead variants, we tested for gene-by-season interaction in PLINK^64^ among unrelated participants of white British ancestry (n =337,409) using linear regression and an additive genetic model. Interaction analyses were adjusted for age, sex, 10 PCs, genotyping array, and season to determine SNP interaction with season on daytime napping. In addition, for each *locus*, corresponding summary statistics for other self-reported and accelerometer-derived sleep measures were obtained from the Sleep Disorder Knowledge Portal (http://sleepdisordergenetics.org/). As earlier UK Biobank GWASs were restricted to the Haplotype Reference Consortium (HRC) imputed variants, if the lead signal was unavailable, a proxy SNP (*r*^2^>0.8) was used instead.

### 23andMe, Inc. replication

23andMe, Inc. is a personal genetics company. DNA extraction and genotyping were performed on saliva samples by National Genetics Institute, a CLIA licensed clinical laboratory and a subsidiary of Laboratory Corporation of America. Samples were genotyped on one of five genotyping platforms. Samples that failed to reach 98.5% call rate were re-analyzed. A single unified imputation reference panel was created by combining the May 2015 release of the 1000 Genomes Phase 3 haplotypes^65^ with the UK10K imputation reference panel^66^. For each chromosome, Minimac3^67^ was used to impute the reference panels against each other, reporting the best-guess genotype at each site. Ancestry was determined through an analysis of local ancestry^68^. A principal component analysis was performed independently for each ancestry, using ∼65,000 high quality genotyped variants present in all five genotyping platforms. In addition, a maximal set of unrelated individuals was chosen for each analysis using a segmental identity-by-descent estimation algorithm. All individuals included in the analyses provided informed consent and answered surveys online according to human subject protocol, which was reviewed and approved by Ethical & Independent Review Services, a private institutional review board (http://www.eandireview.com).

For the present daytime napping replication, we restricted analyses to 541,333 participants of European ancestry with survey responses to a question on frequency of daytime napping. Participants were asked, “How many days per week do you take naps during the day? (15 minutes or more)” with a response on a continuous scale. Responses in days per week were scaled to ‘Never/Rarely’ if 0 or 1 (*n* =267,271), ‘Sometimes’ if 2 to 5 (*n* =232,868), and ‘Usually’ if 6 or 7 (*n* =41,194) to more closely resemble the UK Biobank categories. Replication for the 123 daytime napping loci or proxy for lead SNP (r^2^ >0.80) were generated through linear regression (using an additive model) of the phenotype against the genotype, adjusting for age, sex, the first four principal components and a categorical variable representing genotyping platform. Furthermore, meta-analysis of UK Biobank and 23andMe associations for the daytime napping loci was performed using METAL^69^ by weighting effect size estimates using the inverse of the corresponding standard errors squared (version released March 25 2011).

### Colocalization

To identify genomic regions which harbor causal variants influencing multiple sleep traits, we conducted multi-trait colocalization using the Hypothesis Prioritization Colocalization (HyPrColoc) package^27^. This package performs multi-trait colocalization using a computationally efficient algorithm that facilitates colocalization of large numbers of traits. To identify clusters of colocalized traits, we implemented the branch and bound divisive clustering algorithm using GWAS summary statistics for the following sleep traits in the UK Biobank: sleep duration (*n* =446,118)^25^, insomnia symptoms (*n* =129,270 cases / 108,357 controls)^24^, chronotype (*n* =449,734)^22^, snoring (*n* =421,466), ease of awakening (*n* =451,872), and daytime sleepiness (*n* =452,071). Although these GWAS were conducted in the UK Biobank, the algorithm is robust to inclusion of studies with overlapping participants^27^. Colocalization analysis was performed in pre-defined, approximately independent LD blocks across the genome (1.6 Mb on average)^70^. We used the default variant-level prior probability of a SNP associated with a trait of p_1_ = 1×10^−4^ (prior probability of a SNP being associated with one trait) and y =0.98 (1 - prior probability of a SNP being associated with an additional trait given that the SNP is associated with at least 1 other trait). With these settings, 1 in 200,000 variants are expected to be causal for two traits. Consistent with prior work^27^, we conservatively set both the regional and alignment probabilities to 0.80 so that a cluster of colocalized traits would only be identified if P_R_P_A_ > 0.64. The outputs from the algorithm include: i) colocalized traits, ii) the posterior probability of colocalization, iii) the regional association probability (a measure of degree of shared association, analogous to a phenome-wide association study), iv) the candidate causal variant, and v) the proportion of the posterior probability of colocalization explained by the genetic variant (interpreted as a multi-trait fine-mapping probability). We report loci with posterior probability (pp) for colocalization >0.7, as this cutoff correspond to a false discovery rate <5%^27^.

We performed two additional colocalization analyses. Using summary statistics from a meta-analysis (n ∼700,000) of UK Biobank and the GIANT consortium ^71^, we performed genome-wide colocalization of naps with BMI. To link gene expression to the naps associations, we performed colocalization for all genes located within 1MB of the top signals identified in the naps GWAS. We used summary statistics for expression quantitative trait loci (eQTL) associations identified in the Genotype-Tissue Expression project v7^72^. We prioritized gene expression in the frontal cortex, which was identified by FUMA analysis of GTEX v7 to be the most highly enriched tissue for the naps signals.

### Conditional analysis

To determine independence of the daytime sleepiness and napping association signals at the *HCRTR2* locus, we applied the GCTA COJO algorithm to conduct conditional analyses^73^. We used the UK Biobank sample as the LD reference panel and considered a 1Mb window surrounding the lead *HCRTR2* SNP in the napping GWAS (position 6:55142337). We conditioned on rs2653349 using the --cojo-cond function.

### Bayesian nonnegative matrix factorization (bNMF) clustering and association

We applied the bNMF clustering algorithm^39,74,75^ with the aim of collapsing identified naps *loci* into subgroups of variants based on patterns of association with other sleep traits. The inputs for the bNMF algorithm were the set of the 122 naps GWAS signals (rs10639111 was not included due to missing proxy SNP in association analyses for other sleep traits) oriented to naps-increasing alleles and corresponding association statistics for 17 self-reported and accelerometer-derived sleep traits from the UK Biobank. We generated standardized effect sizes for variant trait associations from GWAS by dividing the estimated regression coefficient by the standard error, using the UK Biobank summary statistic results (variant-trait association matrix [122 by 17]). To enable an inference for latent overlapping modules or clusters embedded in variant-trait associations, we modified the existing bNMF algorithm to explicitly account for both positive and negative associations as was done previously^39,75^.

The defining features of each cluster were determined by the most highly associated traits, which is a natural output of the bNMF approach. bNMF algorithm was performed in R for 1,000 iterations with different initial conditions, and the maximum posterior solution at the most probable number of clusters was selected for downstream analysis (i.e., k=3 for 63% of 1,000 iterations in this analysis, with those same 3 clusters present in an additional 34% of iterations with k=4). The results of the bNMF algorithm provide cluster-specific weights for each variant and trait. Variants and traits defining each cluster were based on a cut-off of weighting of 1.09, which was determined by the optimal threshold to define the beginning of the long-tail of the distribution of cluster’s weights across all clusters (top 5% were considered to be significant).

We compared our clusters from the bNMF algorithm using hierarchical cluster analyses, as was previously conducted for daytime sleepiness^23^. Briefly, the analysis uses the pairwise Euclidean distance between the 122 loci z-scores with the 17 self-reported and accelerometer-derived sleep traits.

### Functional annotations of SNPs and pathway and tissue-enrichment analyses

Functional annotation was carried out in FUMA, with annotations including ANNOVAR^76^. Missense variants of interest were further mapped to protein domains using UniProt^77^. Pathway analysis was conducted using MAGMA^42^ gene-set analysis in FUMA^41^, which uses the full distribution of SNP *P* values and is performed for curated gene sets and GO terms obtained from MsigDB (total of 15,481 pathways). A significance threshold was set after Bonferroni correction accounting for all pathways tested (*P* <0.05/15,481). Gene-based analysis was also performed using Pascal^40^. Pascal gene-set enrichment analysis uses 1,077 pathways from KEGG, REACTOME, BIOCARTA databases, and a significance threshold was set after Bonferroni correction accounting for 1,077 pathways tested (*P* <0.05/1,077). We performed single-cell enrichment analysis (Cell Type function) in FUMA^41^ using our MAGMA gene analysis result and multiple human-specific single-cell expression datasets^78^.

### Genetic correlations with publicly available traits and other sleep traits

Genome-wide genetic correlation analysis were calculated using the implementation of cross-trait LD Score Regression (LDSC)^79–81^ in LDHub^80^. This was conducted using all SNPs from the UK Biobank discovery GWAS found in HapMap3 and included publicly available data from 257 published genome-wide association studies. LDSC estimates genetic correlation between two traits from summary statistics (ranging from -1 to 1) using the fact that the GWAS effect-size estimate for each SNP incorporates effects of all SNPs in LD with that SNP, SNPs with high LD have higher *χ*^2^ statistics than SNPs with low LD, and a similar relationship is observed when single study test statistics are replaced with the product of z-scores from two studies of traits with some correlation. Significance was considered at the Bonferroni correction for all tests performed (*P* <0.05/257 tests). In addition to publicly available summary statistics from LDHub, we also used publicly available summary statistics from earlier UK Biobank GWASs for self-reported and accelerometer-derived sleep traits from the Sleep Disorder Knowledge Portal (http://sleepdisordergenetics.org/) and computed genome-wide genetic correlations using LDSC^79–81^. Finally, we calculated genetic correlations between the sex-specific GWAS to determine the similarity in male and female genetic architecture.

### Phenome-wide association study in the Partners Biobank

The Partners Biobank is a hospital-based cohort study from the Partners HealthCare hospitals in Boston, MA with electronic health record (EHR) and genetic data. Recruitment for the Partners Biobank launched in 2010 and is active at participating clinics at Brigham and Women’s Hospital, Massachusetts General Hospital, Spaulding Rehabilitation Hospital, Faulkner Hospital, McLean Hospital, Newton-Wellesley Hospital, and North Short Medical Center. All patients provided consent upon enrollment and the study protocol was approved by Partners Healthcare Institutional Review Board. To date (07/2019), a total of 104,965 subjects were consented.

Genomic data for 30,683 participants were generated with the Illumina Multi-Ethnic Genotyping Array. The genotyping data were harmonized, and quality controlled with a three-step protocol, including two stages of genetic variant removal and an intermediate stage of sample exclusion. The exclusion criteria for variants were: 1) missing call rate ≥0.05, 2) MAF <0.001, and 3) deviation from Hardy-Weinberg equilibrium (*P* <10^−6^). The exclusion criteria for samples were: 1) sex discordances between the reported and genetically predicted sex, 2) missing call rates per sample ≥0.02, 3) subject relatedness (pairs with estimated identity-by-descent ≥0.125, from which we removed the individual with the highest proportion of missingness), and 4) population structure showing more than four standard deviations within the distribution of the study population, according to the first four principal components. Phasing was performed with SHAPEIT2^82^ and then then imputations were performed with the Haplotype Reference Consortium Panel^83^ using the Michigan Imputation Server^67^. Written consent was provided by all study participants. Approval for analysis of Biobank data was obtained by Partners IRB, protocol #2018P002276.

Participant ancestry was determined using TRACE^84^ and the Human Genome Diversity Project (HGDP)^85^ as a reference panel. Principal component analysis outliers were determined by using a principal component analysis projection of the study samples onto the HGDP reference samples, and subsequently excluded from analysis. To correct for population stratification, we computed principal components using TRACE^84^ in the subset with genetically European ancestry. Furthermore, sample relatedness was determined using PLINK^64^, and subsequently one sample from each related pair was excluded.

In aggregate, participants had a total of 7,422,726 ICD-9 and ICD-10 diagnostic codes corresponding to 784,878 instances of phecodes with at least 2 distinct diagnostic codes. The most prevalent codes were 401.1 (essential hypertension: *n* =11,397 cases) and 745 (pain in joint: *n* =10,333 cases). A total of 951 distinct phecodes had at least 100 cases in the biobank.

We generated a genome-wide polygenic score (GPS) for each individual by summing naps-increasing risk alleles across the genome, each weighted by the beta estimate for that allele from the discovery GWAS, using PRSice^86^. Of the 13,304,132 SNPs, 18,310 duplicated variants and 1,856,569 ambiguous variants were excluded, and a total of 11,429,253 SNPs remained. At each site, clumped SNPs based on association *P* value (the variant with the smallest *P* value within a 250kb range) was retained and all those in linkage disequilibrium, *r*^2^ >0.1, were removed. Following LD clumping, the GPS included 995,188 SNPs.

A total of 20,054,591 physician diagnoses were obtained for genotyped participants (*n* =30,683) as determined from EHR. Same-day duplicated diagnoses (*n* =8,265,731), non-ICD 9/10 codes (*n* =466,866), codes from participants of non-European ancestry (*n* =2,968,741) were removed, and a total of 8,353,253 ICD-9/10 diagnoses were kept in the analysis. Similar ICD-9 and ICD-10 were consolidated and then further collapsed to 1,857 phecodes based on clinical similarity, as determined previously^87^. A total of 88.9% of the 8,353,253 ICD-9/10 codes mapped to a phecode. Participants with at least 2 codes for a specific phecode were considered cases for that respective category, whereas participants with no relevant code for that category were considered controls. Codes with at least 100 prevalent cases were kept in the analysis.

The association between the daytime napping GPS and each of 951 disease code was tested using logistic regression with adjustments for age, sex, genotyping array, and 5 principal components, using the PheWAS R package^88^. Phenome-wide significance was considered at the Bonferroni threshold for 951 tested diseases outcomes and a less stringent FDR correction.

### Daytime napping and cluster-specific polygenic score associations with cardiometabolic and sleep traits

We tested associations between daytime napping polygenic scores comprised of all variants (123 loci) and sub-scores restricted to cluster-specific variants (3 clusters) with a range of cardiometabolic traits using publicly available data (listed in **Supplementary Table 21**) and other sleep traits (for cluster-specific polygenic scores only) using data from the Sleep Disorder Knowledge Portal. We generated weighted polygenic scores calculated by summing the products of the daytime napping-increasing allele SNP multiplied by the scaled effect from the discovery GWAS using the GTX package in R^89^. Results are effect estimates per additional effect allele for more daytime napping.

### Mendelian randomization

Mendelian randomization (MR) can be conceptualized as a naturally randomized experiment whereby individuals are randomized to more or less liability for an exposure on the basis of their inherited genetic variation. This approach rests on the random assortment of alleles at gametogenesis, which substantially reduces the burden of confounding, and eliminates the potential for reverse causality of outcomes on genotype. We created a genetic instrument from the lead daytime napping variants. To avoid collider bias, we did not use napping GWAS summary statistics that were adjusted or conditioned for any other variables (e.g. GWAS in the subjectively healthy population). These variants were further clumped at a between-SNP *r*^2^ <0.01. To facilitate analyses, we utilized the TwoSampleMR package^90^ to extract and harmonize data from outcome GWAS on a range of cardiometabolic traits of interest. For all cardiometabolic traits, we utilized two-sample MR, where the outcome GWAS did not overlap with the naps GWAS. When variants were not in the outcome dataset, we identified variants in linkage disequilibrium with the top variant at *r*^2^ >0.80 using the 1000G European reference data integrated into MRBase. Datasets were harmonized to match effect and reference alleles, and we attempted to match strand ambiguous alleles by allele identity and frequency when possible (MAF >0.42). An analogous approach was taken for reverse MR of adiposity measures (waist circumference, waist-hip-ratio adjusted for BMI, BMI) on daytime napping.

In the case of systolic blood pressure (SBP) and diastolic blood pressure (DBP), for which independent summary statistics are not readily available, we undertook a split-sample MR approach^91^ whereby we randomly split the UK Biobank sample of unrelated participants of White British ancestry into two subsets. We then re-estimated genetic associations of napping with the top variants identified in the discovery GWAS within each subset, as well as the association of those variants with SBP and DBP within each subset. In order to reduce regression dilution bias, SBP and DBP were averaged over two measurements and adjusted for self-reported antihypertensive use as done in prior GWAS^92^. We then ran MR utilizing exposure and outcome associations measured in different strata (e.g. napping associations in stratum 1 on diastolic blood pressure in stratum 2). MR effect estimates of daytime napping on blood pressure were combined across the two estimates using fixed effects meta-analysis, and standardized using the sample standard deviations for SBP (11.25 mmHg) and DBP (20.65 mmHg).

After data harmonization, we used inverse variance weighted (IVW) random-effects meta-analysis as the main analytic approach. To account for multiple comparisons, we used a conservative Bonferonni-adjusted alpha threshold (0.05/19=0.0026). As the IVW approach assumes no unbalanced horizontal pleiotropy, we utilized a range of sensitivity analyses robust to violations of this assumption: MR Egger^93^, the simple and weighted median^94^, and MR-PRESSO^95^. MR Egger models a pleiotropy parameter by fitting an intercept term and adjusts the causal estimates accordingly. Estimation of this additional parameter greatly reduces power in the Egger regression. The median estimators yield valid causal effects provided that less than 50% of the information comes from invalid instrumental variables. We considered consistent effects across multiple methods to strengthen causal evidence.

### *HCRTR1* and *HCRTR2* PheWAS

To assess whether missense variants in *HCRTR1* and *HCRTR2* (rs2271933 and rs2653349) associated with cardiovascular outcomes and risk factors, we extracted variant associations from the largest available GWAS for these phenotypes (**Supplementary Table 24**). As a broader investigation, we used data from a phenome-wide association study of 1,402 ICD-code based phenotypes in UK Biobank^46^, accessed through the following web browser: http://pheweb.sph.umich.edu/SAIGE-UKB/ (**Supplementary Table 25**).

## Data Availability

Summary GWAS statistics are publicly available at The Sleep Disorder Knowledge Portal webpage: www. http://sleepdisordergenetics.org/.

## Acknowledgements and Funding

This research has been conducted using the UK Biobank Resource (application 6818). We would like to thank the participants and researchers from the UK Biobank who contributed or collected data. We would also like to thank the research participants and employees of 23andMe for making this work possible. Members of the 23andMe Research Team include: Michelle Agee, Stella Aslibekyan, Adam Auton, Robert K. Bell, Katarzyna Bryc, Sarah K. Clark, Sarah L. Elson, Kipper Fletez-Brant, Pierre Fontanillas, Nicholas A. Furlotte, Pooja M. Gandhi, Karl Heilbron, Barry Hicks, David A. Hinds, Karen E. Huber, Ethan M. Jewett, Yunxuan Jiang, Aaron Kleinman, Keng-Han Lin, Nadia K. Litterman, Marie K. Luff, Jennifer C. McCreight, Matthew H. McIntyre, Kimberly F. McManus, Joanna L. Mountain, Sahar V. Mozaffari, Priyanka Nandakumar, Elizabeth S. Noblin, Carrie A.M. Northover, Jared O’Connell, Aaron A. Petrakovitz, Steven J. Pitts, G. David Poznik, J. Fah Sathirapongsasuti, Anjali J. Shastri, Janie F. Shelton, Suyash Shringarpure, Chao Tian, Joyce Y. Tung, Robert J. Tunney, Vladimir Vacic, Xin Wang, Amir S. Zare. This work is supported by grants NIH-F32DK102323, NIH-4T32HL007901, NIH-R01DK107859, NIH-R35HL135818, NIH-K23DK114551 (MSU), MGH Research Scholar Fund, Academy of Finland #309643 (HMO) and Medical Research Council grant: MR/M005070/1. This work has been supported in part by The Spanish Government of Investigation, Development and Innovation (SAF2017-84135-R) including FEDER co-funding; The Autonomous Community of the Region of Murcia through the Seneca Foundation (20795/PI/18) and NIDDK R01DK105072 granted to M. Garaulet. The MEGASTROKE project received funding from sources specified at http://www.megastroke.org/acknowledgments.html.

## Author Contributions

The study was designed by HSD, ID, MG, and RS. HSD, ID, JML, YH, MSU, HM, HMO, SEJ, JK, ARW, MNW, SA, MG, and RS participated in acquisition, analysis and/or interpretation of data. HSD, ID, MG, and RS wrote the manuscript and all co-authors reviewed and edited the manuscript, before approving its submission. RS is the guarantor of this work and, as such, had full access to all the data in the study and takes responsibility for the integrity of the data and the accuracy of the data analysis.

## Supplementary Figure Legends

**Supplementary Figure 1**. Q-Q plot for genome-wide association of daytime napping (A). Q-Q plot shows the expected verses observed *P* values from association analysis. Functional consequence of daytime napping variants on genes using FUMA (B).

**Supplementary Figure 2**. Colocalization analysis reveals a shared causal variant increasing FNDC5 gene expression in skeletal muscle and reduced napping liability (A,B) Regional association plots for napping and FNDC5 gene expression at rs2786547and variants within 400kb on chromosome 1. The y-axis shows the - log_10_ P value for each variant in the region, and the x-axis shows the genomic position. Each variant is represented by a filled circle, with the rs2786547 variant colored purple, and nearby variants colored according to degree of linkage disequilibrium (*r*^2^) with rs2786547. The lower panel shows genes located in the displayed region and the blue line corresponds to the recombination rate. (C) Effect of T allele dosage on FNDC5 gene expression in skeletal muscle.

**Supplementary Figure 3**. Clustering of daytime napping variants using Bayesian nonnegative matrix factorization (bNMF) algorithm (A,B) and hierarchal clustering (C).

**Supplementary Figure 4**. Two-sample Mendelian randomization sensitivity analyses for the effect of daytime napping on systolic blood pressure, diastolic blood pressure, and waist circumference.

Abbreviations: DBP: diastolic blood pressure; IVW: inverse-variance weighted; SBP: systolic blood pressure

**Supplementary Figure 5**. Phenome-wide association study (PheWAS) of missense variants in *HCRTR1* and *HCRTR2* on 1,402 ICD-defined outcomes in UK Biobank reveals no phenome-wide significant disease associations.

Each dot represents a phenotype, grouped into phenotype class by color. The y axis represents the -log_10_(p-value), with the dotted horizontal line representing the Bonferonni-corrected p-value threshold for phenome-wide significance.

## References

1. Yang Y, Edery I. Daywake, an Anti-siesta Gene Linked to a Splicing-Based Thermostat from an Adjoining Clock Gene. Curr Biol. 2019;29(10):1728-1734.e4. doi:10.1016/j.cub.2019.04.039

2. Capellini I, Nunn CL, McNamara P, Preston BT, Barton RA. Energetic constraints, not predation, influence the evolution of sleep patterning in mammals. Funct Ecol. 2008;22(5):847–853. doi:10.1111/j.1365-2435.2008.01449.x

3. Gradisar M, Wolfson AR, Harvey AG, Hale L, Rosenberg R, Czeisler CA. The sleep and technology use of Americans: findings from the National Sleep Foundation’s 2011 Sleep in America poll. J Clin Sleep Med. 2013;9(12):1291–1299. doi:10.5664/jcsm.3272

4. Ruggiero JS, Redeker NS. Effects of napping on sleepiness and sleep-related performance deficits in night-shift workers: a systematic review. Biol Res Nurs. 2014;16(2):134–142. doi:10.1177/1099800413476571

5. Hartzler BM. Fatigue on the flight deck: the consequences of sleep loss and the benefits of napping. Accid Anal Prev. 2014;62:309–318. doi:10.1016/j.aap.2013.10.010

6. Vgontzas AN, Pejovic S, Zoumakis E, et al. Daytime napping after a night of sleep loss decreases sleepiness, improves performance, and causes beneficial changes in cortisol and interleukin-6 secretion. Am J Physiol - Endocrinol Metab. 2007;292(1):E253–61. doi:10.1152/ajpendo.00651.2005

7. Cai M, Huang Y, Sun X, He Y, Sun C. Siesta is associated with reduced systolic blood pressure level and decreased prevalence of hypertension in older adults. J Hum Hypertens. 2016;30(3):216–218. doi:10.1038/jhh.2015.70

8. Faraut B, Andrillon T, Vecchierini M-F, Leger D. Napping: A public health issue. From epidemiological to laboratory studies. Sleep Med Rev. 2017;35:85–100. doi:10.1016/j.smrv.2016.09.002

9. Tanabe N, Iso H, Seki N, et al. Daytime napping and mortality, with a special reference to cardiovascular disease: the JACC study. Int J Epidemiol. 2010;39(1):233–243. doi:10.1093/ije/dyp327

10. Stone KL, Ewing SK, Ancoli-Israel S, et al. Self-reported sleep and nap habits and risk of mortality in a large cohort of older women. J Am Geriatr Soc. 2009;57(4):604–611. doi:10.1111/j.1532-5415.2008.02171.x

11. Lin D, Sun K, Li F, et al. Association between habitual daytime napping and metabolic syndrome: a population-based study. Metabolism. 2014;63(12):1520–1527. doi:10.1016/j.metabol.2014.08.005

12. Bursztyn M, Ginsberg G, Hammerman-Rozenberg R, Stessman J. The siesta in the elderly: risk factor for mortality? Arch Intern Med. 1999;159(14):1582-1586. http://www.ncbi.nlm.nih.gov/pubmed/10421281. Accessed August 7, 2019.

13. Yamada T, Shojima N, Yamauchi T, Kadowaki T. J-curve relation between daytime nap duration and type 2 diabetes or metabolic syndrome: A doseresponse meta-analysis. Sci Rep. 2016;6:38075. doi:10.1038/srep38075

14. Celis-Morales C, Lyall DM, Guo Y, et al. Sleep characteristics modify the association of genetic predisposition with obesity and anthropometric measurements in 119,679 UK Biobank participants. Am J Clin Nutr. 2017;105(4):980–990. doi:10.3945/ajcn.116.147231

15. Sayón-Orea C, Bes-Rastrollo M, Carlos S, Beunza JJ, Basterra-Gortari FJ, Martínez-González MA. Association between Sleeping Hours and Siesta and the Risk of Obesity: The SUN Mediterranean Cohort. Obes Facts. 2013;6(4):337–347. doi:10.1159/000354746

16. Lopez-Minguez J, Morosoli JJ, Madrid JA, Garaulet M, Ordoñana JR. Heritability of siesta and night-time sleep as continuously assessed by a circadian-related integrated measure. Sci Rep. 2017;7(1):12340. doi:10.1038/s41598-017-12460-x

17. Spada J, Scholz M, Kirsten H, et al. Genome-wide association analysis of actigraphic sleep phenotypes in the LIFE Adult Study. J Sleep Res. 2016;25(6):690–701. doi:10.1111/jsr.12421

18. Jansen PR, Watanabe K, Stringer S, et al. Genome-wide analysis of insomnia in 1,331,010 individuals identifies new risk loci and functional pathways. Nat Genet. 2019;51(3):394–403. doi:10.1038/s41588-018-0333-3

19. Jones SE, van Hees VT, Mazzotti DR, et al. Genetic studies of accelerometer-based sleep measures yield new insights into human sleep behaviour. Nat Commun. 2019;10(1). doi:10.1038/s41467-019-09576-1

20. Vgontzas AN, Bixler EO, Tan TL, Kantner D, Martin LF, Kales A. Obesity without sleep apnea is associated with daytime sleepiness. Arch Intern Med. 1998;158(12):1333–1337. doi:10.1001/archinte.158.12.1333

21. Isaac RE, Li C, Leedale AE, Shirras AD. Drosophila male sex peptide inhibits siesta sleep and promotes locomotor activity in the post-mated female. Proceedings Biol Sci. 2010;277(1678):65–70. doi:10.1098/rspb.2009.1236

22. Jones SE, Lane JM, Wood AR, et al. Genome-wide association analyses of chronotype in 697,828 individuals provides insights into circadian rhythms. Nat Commun. 2019;10(1):343. doi:10.1038/s41467-018-08259-7

23. Wang H, Lane JM, Jones SE, et al. Genome-wide association analysis of selfreported daytime sleepiness identifies 42 loci that suggest biological subtypes. Nat Commun. 2019;10(1):3503. doi:10.1038/s41467-019-11456-7

24. Lane JM, Jones SE, Dashti HS, et al. Biological and clinical insights from genetics of insomnia symptoms. Nat Genet. 2019;51(3):387–393. doi:10.1038/s41588-019-0361-7

25. Dashti HS, Jones SE, Wood AR, et al. Genome-wide association study identifies genetic loci for self-reported habitual sleep duration supported by accelerometer-derived estimates. Nat Commun. 2019;10(1):1100. doi:10.1038/s41467-019-08917-4

26. Bulik-Sullivan BK, Loh P-R, Finucane HK, et al. LD Score regression distinguishes confounding from polygenicity in genome-wide association studies. Nat Genet. 2015;47(3):291–295. doi:10.1038/ng.3211

27. Foley CN, Staley JR, Breen PG, et al. A fast and efficient colocalization algorithm for identifying shared genetic risk factors across multiple traits. bioRxiv. 2019;44(0):592238. doi:10.1101/592238

28. Cvetkovic-Lopes V, Eggermann E, Uschakov A, et al. Rat Hypocretin/Orexin Neurons Are Maintained in a Depolarized State by TRPC Channels. Bartell PA, ed. PLoS One. 2010;5(12):e15673. doi:10.1371/journal.pone.0015673

29. Chiu CN, Rihel J, Lee DA, et al. A Zebrafish Genetic Screen Identifies Neuromedin U as a Regulator of Sleep/Wake States. Neuron. 2016;89(4):842–856. doi:10.1016/j.neuron.2016.01.007

30. Woods IG, Schoppik D, Shi VJ, et al. Neuropeptidergic signaling partitions arousal behaviors in zebrafish. J Neurosci. 2014;34(9):3142–3160. doi:10.1523/JNEUROSCI.3529-13.2014

31. Hardaway JA, Halladay LR, Mazzone CM, et al. Central Amygdala Prepronociceptin-Expressing Neurons Mediate Palatable Food Consumption and Reward. Neuron. 2019;102(5):1037-1052.e7. doi:10.1016/j.neuron.2019.03.037

32. Lane JM, Liang J, Vlasac I, et al. Genome-wide association analyses of sleep disturbance traits identify new loci and highlight shared genetics with neuropsychiatric and metabolic traits. Nat Genet. 2017;49(2):274–281. doi:10.1038/ng.3749

33. Foltenyi K, Greenspan RJ, Newport JW. Activation of EGFR and ERK by rhomboid signaling regulates the consolidation and maintenance of sleep in Drosophila. Nat Neurosci. 2007;10(9):1160–1167. doi:10.1038/nn1957

34. Lee DA, Liu J, Hong Y, et al. Evolutionarily conserved regulation of sleep by epidermal growth factor receptor signaling. Sci Adv. 2019;5(11). doi:10.1126/sciadv.aax4249

35. Dougherty MK, Ritt DA, Zhou M, et al. KSR2 is a calcineurin substrate that promotes ERK cascade activation in response to calcium signals. Mol Cell. 2009;34(6):652–662. doi:10.1016/j.molcel.2009.06.001

36. Chen C, Xu M, Anantaprakorn Y, Rosing M, Stanewsky R. nocte Is Required for Integrating Light and Temperature Inputs in Circadian Clock Neurons of Drosophila. Curr Biol. 2018;28(10):1595-1605.e3. doi:10.1016/j.cub.2018.04.001

37. Ardlie KG, DeLuca DS, Segrè A V., et al. The Genotype-Tissue Expression (GTEx) pilot analysis: Multitissue gene regulation in humans. Science (80-). 2015;348(6235):648–660. doi:10.1126/science.1262110

38. Natalicchio A, Marrano N, Biondi G, et al. Irisin increases the expression of anorexigenic and neurotrophic genes in mouse brain. Diabetes Metab Res Rev. 2019;36(3). doi:10.1002/dmrr.3238

39. Udler MS, Kim J, von Grotthuss M, et al. Type 2 diabetes genetic loci informed by multi-trait associations point to disease mechanisms and subtypes: A soft clustering analysis. Langenberg C, ed. PLoS Med. 2018;15(9):e1002654. doi:10.1371/journal.pmed.1002654

40. Lamparter D, Marbach D, Rueedi R, Kutalik Z, Bergmann S. Fast and Rigorous Computation of Gene and Pathway Scores from SNP-Based Summary Statistics. Listgarten J, ed. PLoS Comput Biol. 2016;12(1):e1004714. doi:10.1371/journal.pcbi.1004714

41. Watanabe K, Taskesen E, van Bochoven A, Posthuma D. Functional mapping and annotation of genetic associations with FUMA. Nat Commun. 2017;8(1):1826. doi:10.1038/s41467-017-01261-5

42. de Leeuw CA, Mooij JM, Heskes T, Posthuma D. MAGMA: generalized gene-set analysis of GWAS data. PLoS Comput Biol. 2015;11(4):e1004219. doi:10.1371/journal.pcbi.1004219

43. Karlson E, Boutin N, Hoffnagle A, Allen N. Building the Partners HealthCare Biobank at Partners Personalized Medicine: Informed Consent, Return of Research Results, Recruitment Lessons and Operational Considerations. J Pers Med. 2016;6(1):2. doi:10.3390/jpm6010002

44. Dashti HS, Redline S, Saxena R. Polygenic risk score identifies associations between sleep duration and diseases determined from an electronic medical record biobank. Sleep. December 2018. doi:10.1093/sleep/zsy247

45. McAlpine CS, Kiss MG, Rattik S, et al. Sleep modulates haematopoiesis and protects against atherosclerosis. Nature. 2019;566(7744):383–387. doi:10.1038/s41586-019-0948-2

46. Zhou W, Nielsen JB, Fritsche LG, et al. Efficiently controlling for case-control imbalance and sample relatedness in large-scale genetic association studies. Nat Genet. 2018;50(9):1335–1341. doi:10.1038/s41588-018-0184-y

47. Mieda M. The roles of orexins in sleep/wake regulation. Neurosci Res. 2017;118:56–65. doi:10.1016/j.neures.2017.03.015

48. Allada R, Cirelli C, Sehgal A. Molecular Mechanisms of Sleep Homeostasis in Flies and Mammals. Cold Spring Harb Perspect Biol. 2017;9(8):a027730. doi:10.1101/cshperspect.a027730

49. Buxton OM, Lee S, Marino M, Beverly C, Almeida DM, Berkman L. Sleep health and predicted cardiometabolic risk scores in employed adults from two industries. J Clin Sleep Med. 2018;14(3):371–383. doi:10.5664/jcsm.6980

50. Panossian LA, Veasey SC. Daytime Sleepiness in Obesity: Mechanisms Beyond Obstructive Sleep Apnea—A Review. Sleep. 2012;35(5):605–615. doi:10.5665/sleep.1812

51. Ramos AR, Weng J, Wallace DM, et al. Sleep Patterns and Hypertension Using Actigraphy in the Hispanic Community Health Study/Study of Latinos. Chest. 2018;153(1):87–93. doi:10.1016/j.chest.2017.09.028

52. Cheungpasitporn W, Thongprayoon C, Srivali N, et al. The effects of napping on the risk of hypertension: a systematic review and meta-analysis. J Evid Based Med. 2016;9(4):205–212. doi:10.1111/jebm.12211

53. Cao Z, Shen L, Wu J, et al. The effects of midday nap duration on the risk of hypertension in amiddle-aged and older Chinese population: A preliminary evidence from the Tongji-Dongfeng Cohort Study, China. J Hypertens. 2014;32(10):1993–1998. doi:10.1097/HJH.0000000000000291

54. Stergiou GS, Mastorantonakis SE, Roussias LG. Intraindividual reproducibility of blood pressure surge upon rising after nighttime sleep and siesta. Hypertens Res. 2008;31(10):1859–1864. doi:10.1291/hypres.31.1859

55. Stergiou GS, Mastorantonakis SE, Roussias LG. Morning blood pressure surge: The reliability of different definitions. Hypertens Res. 2008;31(8):1589–1594. doi:10.1291/hypres.31.1589

56. Cappuccio FP, Miller MA. Sleep and Cardio-Metabolic Disease. Vol 19. Current Medicine Group LLC 1; 2017. doi:10.1007/s11886-017-0916-0

57. Jack Bowden. Misconceptions on the use of MR-Egger regression and the evaluation of the InSIDE assumption | International Journal of Epidemiology | Oxford Academic. Int J Epidemiol. 2017;46(6):2097–2099. https://academic.oup.com/ije/article/46/6/2097/4157383. Accessed May 5, 2020.

58. Sudlow C, Gallacher J, Allen N, et al. UK biobank: an open access resource for identifying the causes of a wide range of complex diseases of middle and old age. PLoS Med. 2015;12(3):e1001779. doi:10.1371/journal.pmed.1001779

59. Manousaki D, Mitchell R, Dudding T, et al. Genome-wide Association Study for Vitamin D Levels Reveals 69 Independent Loci. Am J Hum Genet. 2020;106(3):327–337. doi:10.1016/j.ajhg.2020.01.017

60. van Hees VT, Sabia S, Jones SE, et al. Estimating sleep parameters using an accelerometer without sleep diary. Sci Rep. 2018;8(1):12975. doi:10.1038/s41598-018-31266-z

61. Bycroft C, Freeman C, Petkova D, et al. The UK Biobank resource with deep phenotyping and genomic data. Nature. 2018;562(7726):203–209. doi:10.1038/s41586-018-0579-z

62. Loh P-R, Tucker G, Bulik-Sullivan BK, et al. Efficient Bayesian mixed-model analysis increases association power in large cohorts. Nat Genet. 2015;47(3):284–290. doi:10.1038/ng.3190

63. Farh KK-H, Marson A, Zhu J, et al. Genetic and epigenetic fine mapping of causal autoimmune disease variants. Nature. 2015;518(7539):337–343. doi:10.1038/nature13835

64. Purcell S, Neale B, Todd-Brown K, et al. PLINK: a tool set for whole-genome association and population-based linkage analyses. Am J Hum Genet. 2007;81(3):559–575. doi:10.1086/519795

65. Auton A, Abecasis GR, Altshuler DM, et al. A global reference for human genetic variation. Nature. 2015;526(7571):68–74. doi:10.1038/nature15393

66. Walter K, Min JL, Huang J, et al. The UK10K project identifies rare variants in health and disease. Nature. 2015;526(7571):82–89. doi:10.1038/nature14962

67. Das S, Forer L, Schönherr S, et al. Next-generation genotype imputation service and methods. Nat Genet. 2016;48(10):1284–1287. doi:10.1038/ng.3656

68. Durand EY, Do CB, Mountain JL, Macpherson JM. Ancestry Composition: A Novel, Efficient Pipeline for Ancestry Deconvolution. bioRxiv. 2014:010512. doi:10.1101/010512

69. Willer CJ, Li Y, Abecasis GR. METAL: fast and efficient meta-analysis of genomewide association scans. Bioinformatics. 2010;26(17):2190–2191. doi:10.1093/bioinformatics/btq340

70. Berisa T, Pickrell JK. Approximately independent linkage disequilibrium blocks in human populations. Bioinformatics. 2016;32(2):283–285. doi:10.1093/bioinformatics/btv546

71. Yengo L, Sidorenko J, Kemper KE, et al. Meta-analysis of genome-wide association studies for height and body mass index in ∼700 000 individuals of European ancestry. Hum Mol Genet. 2018;27(20):3641–3649. doi:10.1093/hmg/ddy271

72. Aguet F, Brown AA, Castel SE, et al. Genetic effects on gene expression across human tissues. Nature. 2017;550(7675):204–213. doi:10.1038/nature24277

73. Yang J, Ferreira T, Morris AP, et al. Conditional and joint multiple-SNP analysis of GWAS summary statistics identifies additional variants influencing complex traits. Nat Genet. 2012;44(4):369–375. doi:10.1038/ng.2213

74. Tan VYF, Févotte C. Automatic relevance determination in nonnegative matrix factorization with the β-divergence. IEEE Trans Pattern Anal Mach Intell. 2013;35(7):1592–1605. doi:10.1109/TPAMI.2012.240

75. Kim J, Mouw KW, Polak P, et al. Somatic ERCC2 mutations are associated with a distinct genomic signature in urothelial tumors. Nat Genet. 2016;48(6):600–606. doi:10.1038/ng.3557

76. Wang K, Li M, Hakonarson H. ANNOVAR: functional annotation of genetic variants from high-throughput sequencing data. Nucleic Acids Res. 2010;38(16):e164. doi:10.1093/nar/gkq603

77. Bateman A. https:UniProt: a worldwide hub of protein knowledge. Nucleic Acids Res. 2019;47(D1):D506–D515. doi:10.1093/nar/gky1049

78. Watanabe K, Umićević Mirkov M, de Leeuw CA, van den Heuvel MP, Posthuma D. https:Genetic mapping of cell type specificity for complex traits. Nat Commun. 2019;10(1):3222. doi:10.1038/s41467-019-11181-1

79. Finucane HK, Bulik-Sullivan B, Gusev A, et al. Partitioning heritability by functional annotation using genome-wide association summary statistics. Nat Genet. 2015;47(11):1228–1235. doi:10.1038/ng.3404

80. Zheng J, Erzurumluoglu AM, Elsworth BL, et al. LD Hub: a centralized database and web interface to perform LD score regression that maximizes the potential of summary level GWAS data for SNP heritability and genetic correlation analysis. Bioinformatics. 2017;33(2):272–279. doi:10.1093/bioinformatics/btw613

81. Bulik-Sullivan B, Finucane HK, Anttila V, et al. An atlas of genetic correlations across human diseases and traits. Nat Genet. 2015;47(11):1236–1241. doi:10.1038/ng.3406

82. Delaneau O, Zagury J-F, Marchini J. Improved whole-chromosome phasing for disease and population genetic studies. Nat Methods. 2013;10(1):5–6. doi:10.1038/nmeth.2307

83. McCarthy S, Das S, Kretzschmar W, et al. A reference panel of 64,976 haplotypes for genotype imputation. Nat Genet. 2016;48(10):1279–1283. doi:10.1038/ng.3643

84. Wang C, Zhan X, Liang L, Abecasis GR, Lin X. Improved ancestry estimation for both genotyping and sequencing data using projection procrustes analysis and genotype imputation. Am J Hum Genet. 2015;96(6):926–937. doi:10.1016/j.ajhg.2015.04.018

85. Cann HM, de Toma C, Cazes L, et al. A human genome diversity cell line panel. Science. 2002;296(5566):261–262. http://www.ncbi.nlm.nih.gov/pubmed/11954565. Accessed September 13, 2018.

86. Euesden J, Lewis CM, O’Reilly PF. https:PRSice: Polygenic Risk Score software. Bioinformatics. 2015;31(9):1466–1468. doi:10.1093/bioinformatics/btu848

87. Wei W-Q, Bastarache LA, Carroll RJ, et al. Evaluating phecodes, clinical classification software, and ICD-9-CM codes for phenome-wide association studies in the electronic health record. Rzhetsky A, ed. PLoS One. 2017;12(7):e0175508. doi:10.1371/journal.pone.0175508

88. Denny JC, Ritchie MD, Basford MA, et al. PheWAS: demonstrating the feasibility of a phenome-wide scan to discover gene-disease associations. Bioinformatics. 2010;26(9):1205–1210. doi:10.1093/bioinformatics/btq126

89. International Consortium for Blood Pressure Genome-Wide Association Studies GB, Ehret GB, Munroe PB, et al. Genetic variants in novel pathways influence blood pressure and cardiovascular disease risk. Nature. 2011;478(7367):103–109. doi:10.1038/nature10405

90. Hemani G, Zheng J, Elsworth B, et al. The MR-Base platform supports systematic causal inference across the human phenome. Elife. 2018;7. doi:10.7554/eLife.34408

91. Henry A, Katsoulis M, Masi S, et al. The relationship between sleep duration, cognition and dementia: a Mendelian randomization study. Int J Epidemiol. May 2019:1–12. doi:10.1093/ije/dyz071

92. Giri A, Hellwege JN, Keaton JM, et al. Trans-ethnic association study of blood pressure determinants in over 750,000 individuals. Nat Genet. 2019;51(1):51–62. doi:10.1038/s41588-018-0303-9

93. Bowden J, Smith GD, Burgess S. Mendelian randomization with invalid instruments: Effect estimation and bias detection through Egger regression. Int J Epidemiol. 2015;44(2):512–525. doi:10.1093/ije/dyv080

94. Bowden J, Davey Smith G, Haycock PC, Burgess S. Consistent Estimation in Mendelian Randomization with Some Invalid Instruments Using a Weighted Median Estimator. Genet Epidemiol. 2016;40(4):304–314. doi:10.1002/gepi.21965

95. Verbanck M, Chen C-Y, Neale B, Do R. Detection of widespread horizontal pleiotropy in causal relationships inferred from Mendelian randomization between complex traits and diseases. Nat Genet. 2018;50(5):693–698. doi:10.1038/s41588-018-0099-7

96. Lin L, Faraco J, Li R, et al. The sleep disorder canine narcolepsy is caused by a mutation in the hypocretin (orexin) receptor 2 gene. Cell. 1999;98(3):365–376. doi:10.1016/S0092-8674(00)81965-0

